# Equipment-free detection of SARS-CoV-2 and Variants of Concern using Cas13

**DOI:** 10.1101/2021.11.01.21265764

**Authors:** Jon Arizti-Sanz, A’Doriann Bradley, Yibin B. Zhang, Chloe K. Boehm, Catherine A. Freije, Michelle E. Grunberg, Tinna-Solveig F. Kosoko-Thoroddsen, Nicole L. Welch, Priya P. Pillai, Sreekar Mantena, Gaeun Kim, Jessica N. Uwanibe, Oluwagboadurami G. John, Philomena E. Eromon, Gregory Kocher, Robin Gross, Justin S. Lee, Lisa E. Hensley, Christian T. Happi, Jeremy Johnson, Pardis C. Sabeti, Cameron Myhrvold

## Abstract

The COVID-19 pandemic, and the recent rise and widespread transmission of SARS-CoV-2 Variants of Concern (VOCs), have demonstrated the need for ubiquitous nucleic acid testing outside of centralized clinical laboratories. Here, we develop SHINEv2, a Cas13-based nucleic acid diagnostic that combines quick and ambient temperature sample processing and lyophilized reagents to greatly simplify the test procedure and assay distribution. We benchmarked a SHINEv2 assay for SARS-CoV-2 detection against state-of-the-art antigen-capture tests using 96 patient samples, demonstrating 50-fold greater sensitivity and 100% specificity. We designed SHINEv2 assays for discriminating the Alpha, Beta, Gamma and Delta VOCs, which can be read out visually using lateral flow technology. We further demonstrate that our assays can be performed without any equipment in less than 90 minutes. SHINEv2 represents an important advance towards rapid nucleic acid tests that can be performed in any location.

## Introduction

Frequent and widespread testing is critical to prevent and respond to infectious disease outbreaks. For example, large-scale testing to track the prevalence and transmission of severe acute respiratory syndrome coronavirus 2 (SARS-CoV-2) has been essential to manage the ongoing coronavirus disease 2019 (COVID-19) pandemic^1–3^. Ubiquitous and frequent diagnostic testing leads to the rapid identification of new cases and permits the swift treatment or isolation of infected individuals, thereby preventing further viral spread^4^. However, the gold standard for COVID-19 diagnosis, reverse transcription quantitative polymerase chain reaction (RT-qPCR), remains suboptimal for orchestrating this response. RT-qPCR requires specialized equipment and expertise rarely found outside of centralized laboratories. In addition, an insufficient testing infrastructure coupled with reagent shortages and high testing demand have led to slow sample-to-answer times (often of several days) for RT-qPCR^5,6^. Alternative diagnostic technologies that enable rapid and decentralized testing are vital to respond to the current and future pandemics.

Lateral flow antigen-capture tests and isothermal nucleic acid diagnostics with visual readouts represent promising alternatives for SARS-CoV-2 testing outside of centralized laboratories. The effectiveness of such tests in curbing the spread of SARS-CoV-2 has been demonstrated in multiple settings, ranging from nursing homes to whole countries^3,7^. Antigen-capture tests, such as Abbott’s BinaxNow COVID - 19 Antigen Self Tests, are quick and user-friendly but their moderate sensitivity means they can miss potentially infectious individuals^8–10^. Isothermal nucleic acid amplification methods, such as loop-mediated isothermal amplification (LAMP), are more sensitive than antigen-capture tests and operate at a single temperature^11,12^. The deployment of LAMP-based tests has been facilitated by the use of auxiliary devices to eliminate the need for intensive nucleic acid purification and maintain a stable temperature during amplification^13^. However, these devices are often too costly for single-use and difficult to manufacture at the scale required for population-wide distribution, which can limit their utility for frequent and widespread testing^14^. Hence, the continued development and deployment of user-friendly, sensitive and equipment-free diagnostics is key to enhance the public health response to COVID-19.

CRISPR-based diagnostics (CRISPR-Dx) are promising technologies for SARS-CoV-2 testing with minimal equipment requirements. CRISPR-Dx usually combine isothermal nucleic acid amplification methods (often LAMP or recombinase polymerase amplification (RPA)) with a RNA-guided CRISPR-Cas nuclease (usually Cas12 or Cas13) for pathogen detection^15–18^. Detection of an amplified target nucleic acid triggers cleavage of a reporter molecule by the CRISPR-Cas enzyme. This reporter cleavage can be detected by the appearance of visual bands on a lateral flow strip, among other methods^18,19^. Pairing isothermal nucleic acid amplification methods with Cas nuclease detection considerably enhances specificity and sensitivity, but at the expense of increasing assay complexity. Some groups have simplified the workflow by eliminating the need for amplification using more active Cas enzymes, although they rely on custom-built equipment and nucleic acid extraction^20,21^. Several groups have combined multiple enzymatic steps into individual reactions to streamline CRISPR-Dx^22,23^. We previously developed SHINE (<u>S</u>treamlined <u>H</u>ighlighting of <u>I</u>nfections to <u>N</u>avigate <u>E</u>pidemics), henceforth SHINEv1, a diagnostic assay that does not require nucleic acid extractions or custom equipment^23^. However, SHINEv1 involves multiple heating steps, and uses reagent mixes that require cold temperature storage and manual preparation by trained personnel. Further simplifications and technical improvements to CRISPR-Dx would greatly facilitate test distribution and expand their use cases.

Frequent and widespread testing is even more important with the rise of highly transmissible SARS-CoV-2 variants. In recent months, the evolution of SARS-CoV-2 has led to the emergence and sustained transmission of viral variants with sets of mutations that can increase infectivity or reduce neutralization by antibodies, making the virus harder to contain^24^. The World Health Organization (WHO) has designated some of the SARS-CoV-2 variants with increased transmissibility, virulence and/or antigenicity as Variants of Concern (VOCs). Notably, the Alpha, Beta, Gamma and Delta VOCs (designated B.1.1.7, B.1.351, P.1 and B.1.617.2 with the PANGO nomenclature system, respectively) are widely circulating in many countries around the world and have placed unprecedented strain on several healthcare systems^25,26^. Surveillance of SARS-CoV-2 VOCs is vital to inform public health and clinical decisions, such as for prioritizing testing or vaccination in the populations most heavily impacted by the pandemic. Currently, the identification of viral mutations and tracking of circulating SARS-CoV-2 variants is performed by viral genomic sequencing and a small number of mutation-specific RT-qPCR tests^27–29^. However, the lack of infrastructure and expertise have made routine genomic surveillance difficult to perform outside of specialized genomic centers^30^.

CRISPR-Dx are uniquely suited to detect VOCs for routine surveillance applications. In contrast to available point-of-need tests, CRISPR-Dx can easily detect individual SARS-CoV-2 mutations. They do so by measuring relative signal intensities using a set of two crRNAs designed against the variant region^15,19,31^. To date, only a few CRISPR-Dx have been developed for the identification of single or multiple nucleotide substitutions present in various SARS-CoV-2 VOCs^32–35^. However, these tests consist of multiple liquid handling steps or rely on custom-built equipment^32,35^. The development of equipment-free and user-friendly CRISPR-Dx assays to distinguish SARS-CoV-2 VOCs would fill a major unmet need in COVID-19 diagnostics.

Here, we develop SHINEv2, a fast, user-friendly and widely deployable technology for the detection of SARS-CoV-2 and its variants in clinical samples. We improve upon SHINEv1 by incorporating a fast and ambient-temperature sample lysis procedure and enabling the lyophilization of the assay reagents. These improvements considerably reduce assay complexity and facilitate its distribution. We demonstrate that SHINEv2 can detect SARS-CoV-2 RNA in unextracted patient samples with 100% specificity and 50-fold higher sensitivity than cutting-edge antigen-capture tests. Moreover, we designed and validated SHINEv2 assays to discriminate the Alpha, Beta, Gamma and Delta VOCs in clinical samples. Finally, we introduced a Cas12-based human RNase P control, further simplified the lateral flow assay and examined the performance of SHINEv2 at ambient-temperature. These additional advances could expand the use cases of SHINEv2 to virtually any location, including at-home settings.

## Results

### Design of SHINE assay for SARS-CoV-2 detection - S gene assay

We previously developed a SHINEv1 assay to detect SARS-CoV-2 open reading frame 1a (ORF1a) directly from clinical samples^23^. We wondered if the performance of SHINE could be further enhanced through the selection of target sites with more active RPA primers and crRNAs. Using ADAPT^36^, a genomics-informed computational design tool for nucleic acid based diagnostics, on 308,315 SARS-CoV-2 genomes collected before February 2021, we identified RPA primers and 3 crRNAs with high predicted activity in detecting the S gene of SARS-CoV-2. We selected the most active crRNA by measuring Cas13 cleavage activity on synthetic RNA targets without prior amplification (Supplementary Fig. 1) and we optimized the RPA primer concentration to create the S gene assay. We performed bioinformatic analysis to ensure that the crRNA and RPA primer sequences were highly conserved across SARS-CoV-2 genomes. The crRNA sequence was fully conserved in 99.79% of the 308,315 SARS-CoV-2 genomes analysed and the RPA primers were predicted to amplify 99.77% of the genomes. When compared to the ORF1a assay, the S gene assay was found to be 1-2x faster and 10x more sensitive (Supplementary Fig. 2). Therefore, given its increased sensitivity and favorable kinetics, we selected the S gene assay for further development.

### Improving the accessibility of SHINEv1

For SARS-CoV-2 diagnostic testing to occur in virtually any location, improvements were needed to increase user-friendliness and facilitate test distribution. Towards this goal, we developed SHINEv2, which incorporates ambient-temperature sample processing and lyophilized testing reagents, thereby greatly simplifying reagent transport and storage and decreasing the overall complexity of the assay.

To be an effective approach, extraction-free sample processing must inactivate nucleases and infectious viral particles, and must be compatible with the assay, allowing for good diagnostic performance. Previously, we circumvented the need for clinical sample extraction using HUDSON, a nuclease inactivation and viral lysis method that relies on heat and chemical reduction^19,23^.To eliminate HUDSON’s need for heating elements and incubation at two temperatures, we sought to find an ambient temperature alternative for sample processing. FastAmp Viral and Cell Solution for COVID-19 testing (Intact Genomics) functions at ambient temperature and has been reported to be compatible with isothermal nucleic acid amplification methods^37^. Therefore, we investigated whether this lysis reagent was also compatible with Cas13-based detection methods.

We assessed the ability of the FastAmp lysis reagent to inactivate RNases in contrived nasal samples. Our contrived samples consisted of 10% (v/v) healthy nasal fluid in universal transport medium (UTM). After treatment, we added RNaseAlert, a reporter that fluoresces in the presence of RNase activity (Fig. 1a). Without treatment, contrived nasal samples contained high levels of RNases, as indicated by the high fluorescence.

**Fig. 1.**
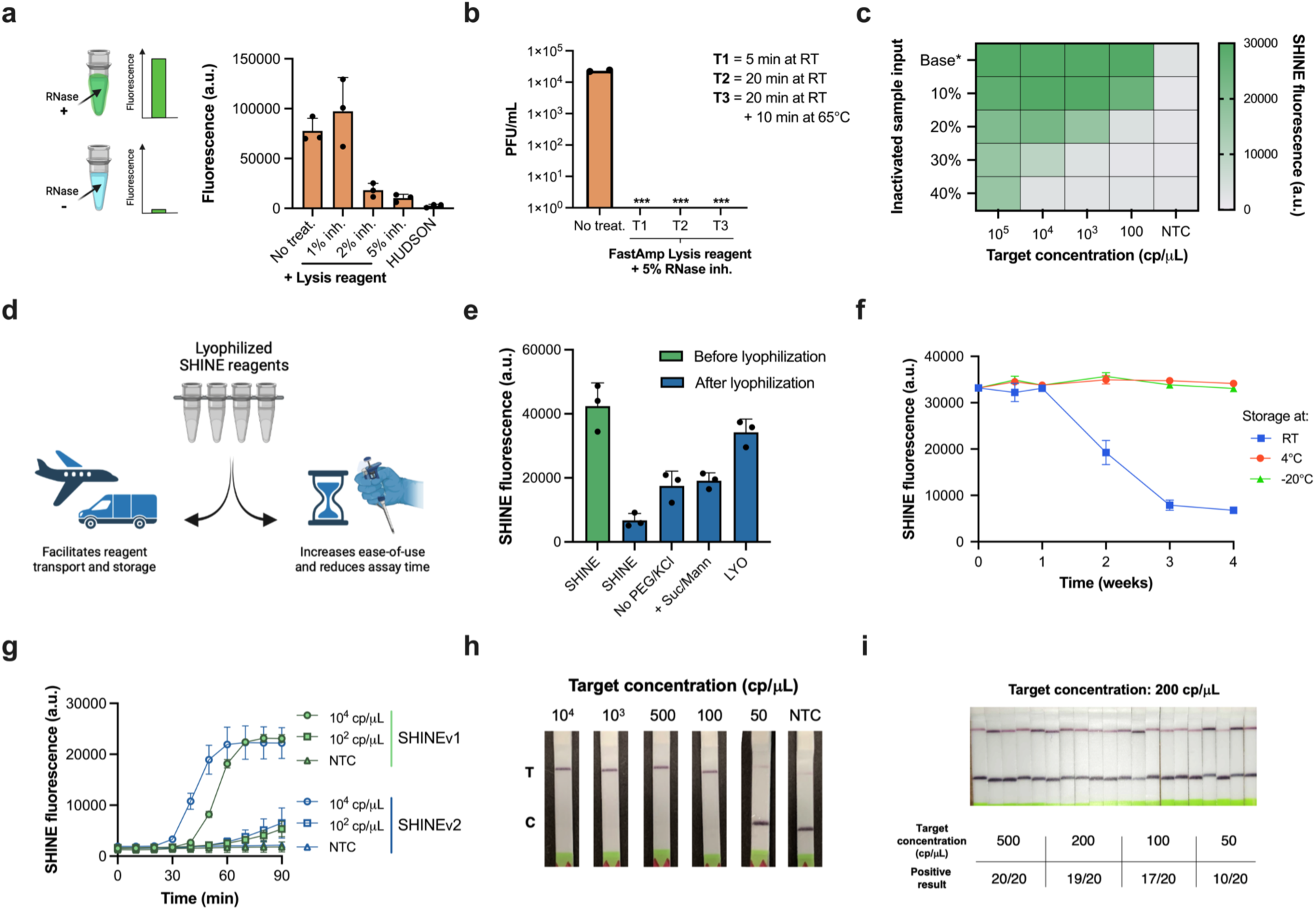
Increasing the ease-of-use and deployability of SHINE. **a**, RNase activity in nasal fluid mixed with universal viral transport medium (UTM) untreated or treated with FastAmp Lysis reagent supplemented with RNase inhibitor (inh.) or treated with HUDSON (a heat- and chemical-treatment). Activity measured using RNaseAlert at room temperature (RT) for 30 minutes. **b**, SARS-CoV-2 seedstock titer without treatment or after being incubated with lysis reagent (+5% RNase inh.) at RT for 5 minutes, 20 minutes or 20 minutes plus 10 minutes at 65°C. ***, infection not detected; PFU, plaque forming units. **c**, SHINE fluorescence with different proportions of inactivated sample input (i.e. FastAmp lysis reagent, RNase inh. and UTM) after a 90 minute incubation. Base*, baseline (i.e. no inactivated sample added). **d**, Schematic of the advantages of lyophilizing SHINE. **e**, SHINE fluorescence on synthetic RNA target (10^4^ copies/μL) before and after lyophilization using different buffers. Fluorescence measured after 90 minutes. For buffer composition, see *Methods*. **f**, SHINE fluorescence after a 90 minute incubation using lyophilized (LYO) reagents stored at RT, 4°C or -20°C over time. Target concentration: 10^4^ copies/μL. **g**, Fluorescence kinetics for SHINEv1 and SHINEv2 using synthetic RNA targets; NTC, no target control. h, Lateral flow detection of SARS-CoV-2 RNA in lysis buffer treated viral seedstocks using SHINEv2, after a 90 minute incubation. C = control band; T = test band. **i**, Determination of analytical limit of detection with 20 replicates of SHINEv2 at different concentrations of SARS-CoV-2 RNA from lysis reagent treated contrived samples. Incubated for 90 minutes. For (**a,e,g**), center = mean and error bars = standard deviation (s.d.) for 3 technical replicates. In **c**, the heatmap values represent the mean for 3 technical replicates. For **f**, center = mean and error bars = s.d. for 3 biological replicates with 3 technical replicates each.

Treatment with FastAmp lysis reagent supplemented with 5% RNase inhibitors decreased the RNase activity by over 85% in contrived nasal samples (Fig. 1a). In addition, the ratio of nasal fluid to UTM in clinical samples derived from nasal swabs is likely to be significantly lower than 10% and therefore, the residual RNase activity in lysed patient samples is expected to be even lower.

Having determined the nuclease inactivation capacity of this lysis solution, we tested its ability to inactivate SARS-CoV-2 viral seedstocks. For each treatment, we quantified the amount of viable virus after two passages using a plaque-based assay (Fig. 1b). Incubation of SARS-CoV-2 with the lysis solution for as little as 5 minutes at room temperature (RT) yielded no discernible plaques, thereby confirming the ability of our lysis solution to swiftly inactivate the virus.

We then examined the compatibility of this lysis solution with good SHINE assay performance. Solution-based sample processing methods often introduce an upper bound of inactivated sample input that can be added, which is dictated by the balance between increased sample volume and decreased assay performance caused by the lysis reagents. Therefore, we supplemented these SHINE reactions with different proportions of inactivated sample inputs (i.e. contrived nasal samples treated with lysis solution) (Fig. 1c). An input of 10% (v/v) inactivated sample into SHINE was able to retain full activity while still doubling the amount of sample added with respect to the previous instantiation of SHINE, which used 5% (v/v) inactivated sample^23^ (Supplementary Fig. 3). Larger input volumes, which would increase the volume of clinical sample added, led to a marked decrease in the sensitivity of SHINE. (Fig. 1C and Supplementary Fig. 3). Thus, our lysis procedure - FastAmp lysis reagent with 5% RNase inhibitor for 5 min - is compatible with SHINE and can effectively inactivate nucleases and SARS-CoV-2 virions, making it ideal for rapid and equipment-free sample processing at ambient temperature.

Lyophilization can facilitate the transport and storage of point-of-care CRISPR-Dx by eliminating the need for a cold chain and increasing the shelf life (Fig. 1d). Lyophilization also simplifies reaction setup, by allowing the full reaction to be easily reconstituted by the end user. Initially, lyophilization with the original SHINE buffer led to a profound drop in activity post-lyophilization (Fig. 1e). We sought to improve the lyophilization process through the addition of non-reducing disaccharides (e.g. sucrose), which can act as stabilizers; the addition of bulking agents (e.g. mannitol) and the removal of potentially destabilizing components, such as polyethylene glycol (PEG) and potassium chloride (KCl). Specifically, the removal of 3.5% PEG-8000 and 60 mM KCl from the original SHINE buffer or the addition of 5% (w/v) sucrose and 150 mM mannitol each moderately increased SHINE’s activity post-lyophilization. With these modifications combined (i.e. LYO buffer), SHINE retained its full activity after lyophilization (Fig. 1e and Supplementary Fig. 4).

Having optimized the lyophilization excipients and procedure, we conducted a month-long experiment to determine the stability of the lyophilized SHINE reagents at different temperatures over time. After being stored at RT, 4°C or -20°C for 4, 7, 14, 21 or 28 days, SHINE pellets were rehydrated and tested on full genome synthetic RNA standards (Twist Bioscience). SHINE pellets retained full activity for >1 week at RT and at least a month at 4°C/-20°C (Fig. 1f). By enabling transport and storage at above-freezing temperatures, lyophilized SHINE greatly simplifies distribution logistics.

Our SHINEv2 technology, which incorporates a new sample lysis procedure and lyophilization into SHINEv1, can sensitively detect nucleic acids with a simple workflow. SHINEv2 substantially reduced the hands-on time and liquid-handling steps needed from the user, from over 45 minutes and 20 pipetting steps in SHINEv1 to less than 10 minutes and 5 user-operations in SHINEv2 (Supplementary Fig. 5). When tested side-by-side with SHINEv1, we found the sensitivity and reaction kinetics of the two methods to be equivalent at multiple target concentrations (Fig. 1g). SHINEv2 detected down to 100 copies per microliter (cp/μL) within 90 minutes, using either a plate-based fluorescence readout or a colorimetric lateral flow based readout (Fig. 1g and 1h). Moreover, we evaluated the impact of transportation and storage on SHINEv2 by comparing its performance over time in a second laboratory in Nigeria. Six weeks after production, and after transportation to a different continent, SHINEv2 was shown to detect down to 100 cp/μL of full genome synthetic RNA standards using a colorimetric lateral flow based readout (Supplementary Fig. 6).

We sought to determine the analytical limit-of-detection (LoD) of SHINEv2 following the guidance released by the Food and Drug Administration (FDA)^38^. Under this guidance, the LoD is defined as the lowest concentration at which at least 19/20 technical replicates are detected. Using SHINEv2 in lateral flow format, we tested 20 replicates of a dilution series of heat-inactivated SARS-CoV-2 viral seedstock in UTM. We determined the analytical LoD of SHINEv2 to be 200 cp/μL (Fig. 1i and Supplementary Fig. 7), which is over an order of magnitude lower than that of state-of-the-art antigen-capture tests and comparable to that of some equipment-based isothermal nucleic acid tests^39,40^. Given the optimal performance of SHINEv2 on contrived SARS-CoV-2 samples, we sought to determine its performance on clinical samples.

### Assessing the clinical performance of SHINEv2 on NP swab samples

We assessed the clinical performance of SHINEv2 on 96 unextracted nasopharyngeal (NP) swab samples from SARS-CoV-2 positive and negative patients, compared to three widely used FDA EUA-authorized diagnostic tests. We first compared the performance of SHINEv2 to the CDC-recommended RT-qPCR protocol for SARS-CoV-2, as the gold-standard reference test ^41^. We also compared SHINEv2 to two state-of-the-art antigen-capture tests: Abbott’s BinaxNow COVID-19 antigen self-test (BinaxNow) and Access Bio’s CareStart COVID-19 antigen test (CareStart)^42,43^. The BinaxNow test, the most widely used rapid test in the United States, represents the cutting-edge in antigen-capture tests. As a second point of comparison, we selected the CareStart test, which has the most similar workflow to SHINEv2 and has comparable performance to BinaxNow.

We performed SHINEv2 testing side-by-side with the RT-qPCR, BinaxNow and CareStart tests (Fig. 2a,b). RT-qPCR detected SARS-CoV-2 RNA in 63 samples out of a total of 96 samples tested, with cycle threshold (C_t_) values ranging from 14.7 to 37.2 (Supplementary Table 1). SHINEv2 detected SARS-CoV-2 RNA in 9/9 RT-qPCR positive samples with C_t_ < 25 (100% sensitivity), 32/34 samples with C_t_ < 30 (94.1% sensitivity), and 34/63 samples with C_t_ < 40 (54.0% sensitivity), (Fig. 2c and Supplementary Fig. 8-10). Notably, our assay correctly identified all positive samples with titers above our analytical LoD of 200 copies/μL and several samples below this LoD. This confirmed that the clinical performance of SHINEv2 is equivalent to its analytical performance.

**Fig. 2.**
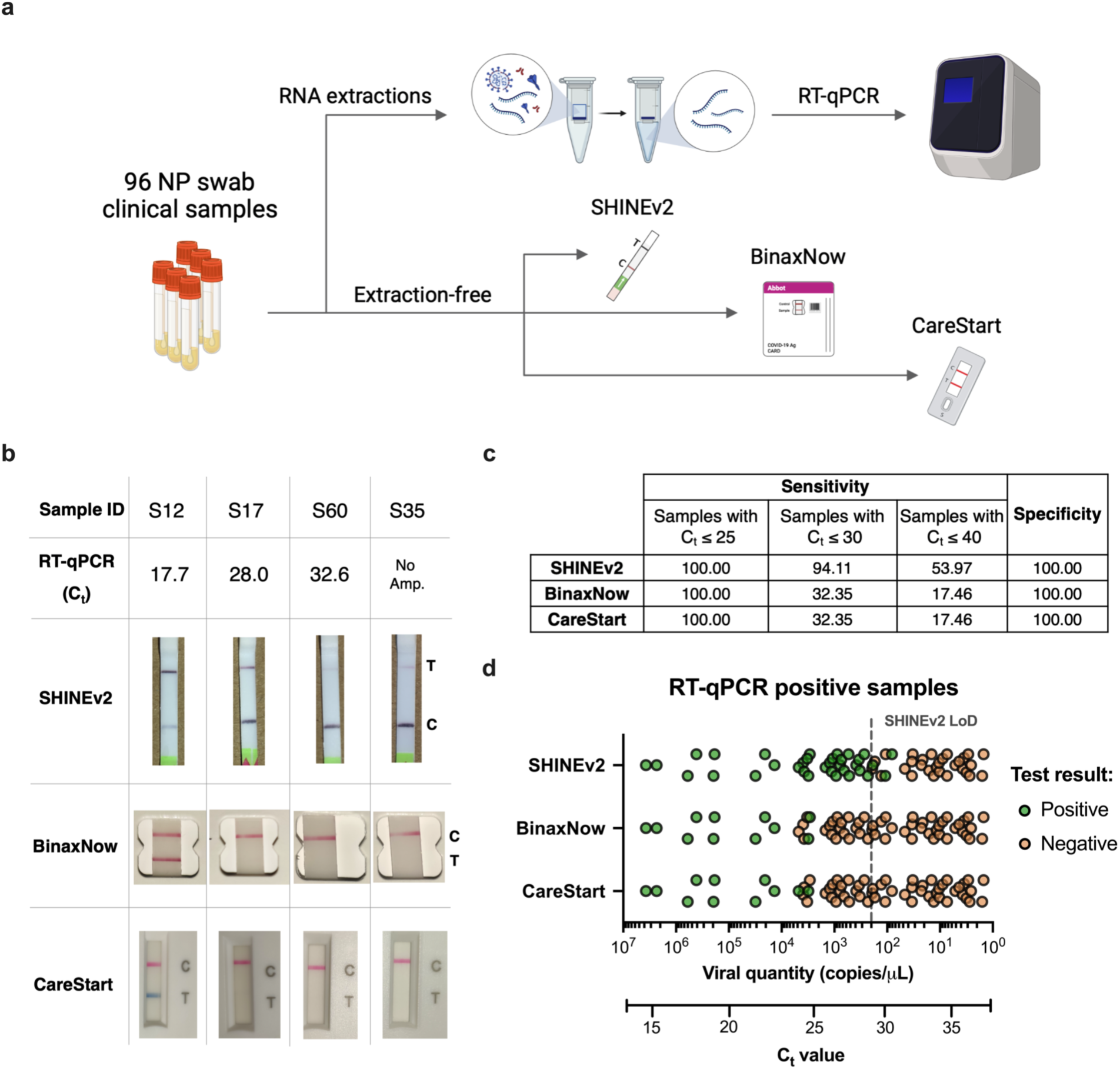
Performance of SHINEv2 on clinical samples. **a,** Schematic of side-by-side clinical sample testing using SHINEv2, BinaxNow, CareStart and RT-qPCR. NP, nasopharyngeal swab. **b,** SHINEv2, BinaxNow and CareStart test results for a subset of clinical NP swab samples with different C_t_ values (CDC EUA N1 RT-qPCR). C = control band; T = test band. No Amp., no amplification detected. For all test results, *see Supplementary Fig. 8-10*. **c,** Side-by-side clinical performance of SHINEv2, BinaxNow and CareStart versus RT-qPCR. **d,** Positive and negative test results for SHINEv2, BinaxNow and CareStart tests for RT-qPCR-positive clinical samples relative to viral RNA concentration and C_t_ value.

Moreover, SHINEv2 was 50 times more sensitive than either antigen-capture test ^42–45^ (Fig. 2d). SHINEv2 excelled at detecting samples with moderate viral loads (25-30 C_t_), thereby allowing the identification of potentially infectious individuals that antigen-capture tests would miss^8–10^. Additionally, SHINEv2, BinaxNow and CareStart were shown to be highly specific, each having no false positive results in any of the 33 RT-qPCR-negative samples (100% specificity) (Fig. 2c). This is consistent with previously reported results for these antigen-capture tests ^42,43^ and is also expected for CRISPR-Dx, given that the RPA primers during amplification and the crRNA during Cas13-based detection provide two layers of specificity^15^. Importantly, the sensitivity and specificity of SHINEv2 on patient samples falls within the recommended range for population-wide screening in settings of reopening ^10,46^.

### Design and testing of SHINEv2 assays for SARS-CoV-2 VOC identification

The emergence of SARS-CoV-2 variants with increased transmissibility and virulence, and the specificity and flexibility of SHINEv2, prompted us to develop assays to detect and distinguish four widely circulating VOCs. Initially, we focused on detecting the 69/70 deletion in the S gene, a hallmark of the Alpha VOC^24,47^. Then, we expanded to the K417N and K417T mutations found in the Beta and Gamma VOCs, respectively^24,47^. Finally, in light of the rampant spread of the Delta VOC in many countries around the world, we sought to rapidly develop new SHINEv2 assays to detect the L452R mutation and 156/157 deletion + R158G mutation present in this variant ^47,48^. We designed sets of crRNAs with differential activities on the reference RNA genome (ancestral target) with respect to the RNA genome containing a given mutation (derived target), or vice versa (Fig. 3a).

**Fig. 3.**
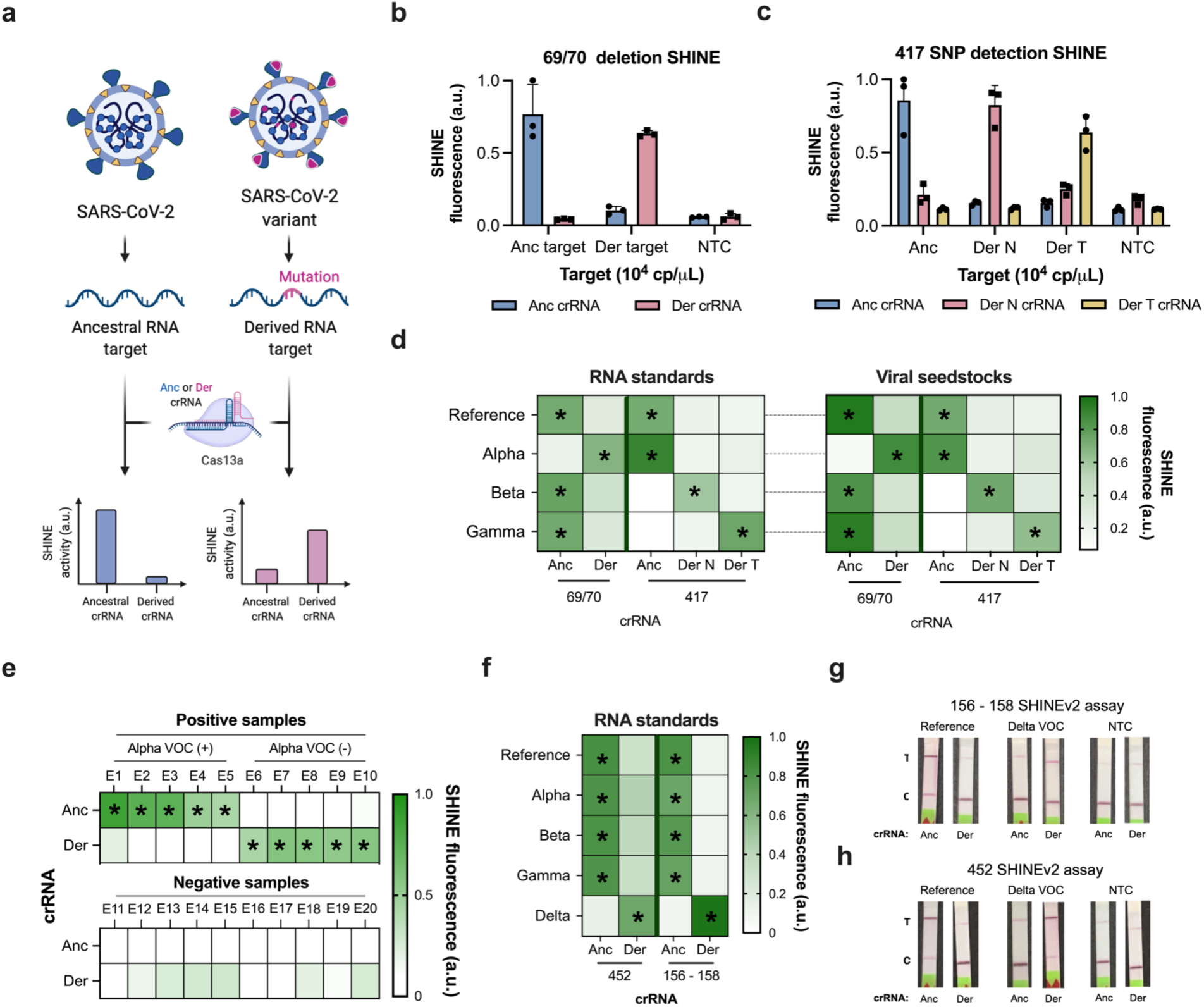
Development of SHINEv2 assays for the detection of SARS-CoV-2 VOCs. **a,** Schematic of Cas13a-based detection of mutations in SARS-CoV-2 using a fluorescent readout; anc, ancestral; der, derived. (**b,c),** SHINE fluorescence of the ancestral and derived crRNAs for the **(b)** 69/70 deletion assay and **(c)** on synthetic RNA targets after 90 minutes; NTC, no target control. **d,** Identification of SARS-CoV-2 VOCs using SHINE fluorescence on full-genome synthetic RNA standards and RNA extracted from viral seedstocks; target RNA concentration: 10^4^ copies/μL. **e,** Mean fluorescence of 69/70 SHINEv2 assay on SARS-CoV-2 RNA extracted from clinical samples, after 90 minutes. **f,** Discrimination of SARS-CoV-2 VOCs using SHINE fluorescence of Delta assays on full-genome synthetic RNA standards, after 90 minutes. Target RNA concentration: 10^4^ genomes/μL. (**g,h),** Colorimetric lateral flow based detection of full-genome synthetic RNA standards using the **(g)** 156 - 158 and **(h)** SHINEv2 assays. SHINEv2 incubation time: 90 minutes. NTC, no target control. T, test line; C, control line. For **b** and **c**, center = mean and error bars = s.d. for 3 technical replicates. In **d,e** and **f**, the heatmap values represent the mean for 3 technical replicates.

For the Alpha VOC, we manually designed a set of 10 ancestral and derived crRNAs to detect the 69/70 deletion. We selected an ancestral crRNA and a derived crRNA based on Cas13 cleavage activity and their ability to discriminate between ancestral and derived RNA targets (Supplementary Fig. 11). The 69/70 deletion falls within the region amplified by the RPA primers from the S gene assay we previously developed (Supplementary Fig. 12). Using these primers, the 69/70 assay successfully detected the 69/70 deletion in synthetic RNA targets (Supplementary Fig. 13). For instance, on the ancestral RNA target, using the ancestral crRNA led to 7x greater fluorescence than the derived crRNA (Fig. 3b). Conversely, on derived RNA target, we observed 5x higher fluorescence with the derived crRNA than with the ancestral crRNA.

Next, we sought to design additional assays for the identification of the Beta and Gamma VOCs. These variants contain distinct single nucleotide polymorphisms (SNPs) at amino acid position 417 in the S gene (i.e. K417N for Beta and K417T for Gamma), none of which are present on the reference genome or the Alpha VOC ^24,47^. However, discriminating between 3 different codons in the same amino acid position using point-of-care diagnostics is remarkably difficult and, to our knowledge, has not been reported to date. We used a computational design technique (see Methods; manuscript *in prep*.) to generate crRNAs that maximize predicted SNP discrimination at position 417, and used ADAPT^36^ to help in designing RPA primers for these crRNAs. After assay optimization, we demonstrated that our 417 assays can differentiate between the three genotypes with high accuracy (Fig. 3c and Supplementary Fig. 14). When tested on synthetic RNA targets, each 417 assay was shown to be at least 3x more active (i.e. higher fluorescence) on its preferred target than on the non-preferred targets (Fig. 3c).

Having demonstrated the sensitivity and specificity of our 69/70 and 417 assays on short synthetic RNA targets, we next tested their performance on full genome synthetic RNA standards and on RNA extracted from SARS-CoV-2 viral seedstocks. In each case, these assays correctly identified the presence or absence of the mutations (Fig. 3d). For the Alpha VOC, the fluorescence signal of the derived crRNA was over 3x higher than that of the ancestral crRNA, indicating the presence of the 69/70 deletion. Conversely, the fluorescence signal for the ancestral crRNA was 1.25-3.5x higher for the reference strain as well as the Beta and Gamma VOCs, which confirmed the absence of the S gene deletion in these cases. Similarly, the genotype of each variant was correctly identified with the 417 assays, based on which crRNA showed the highest activity (Fig 3d). For example, for the Gamma VOC, the derived T crRNA led to the highest fluorescence level, thereby correctly identifying the presence of the K417T SNP in this variant.

Given the promising performance of these assays, we sought to assess their capacity to detect SARS-CoV-2 VOCs in clinical samples. We focused on validating the Alpha VOC assay, the only assay for which we had access to clinical samples, and reasoned that the results obtained could be extended to the other assays. We assessed the clinical performance of the 69/70 assay on RNA extracted from 20 NP swab samples. SHINEv2 correctly identified the presence of the 69/70 deletion in the Alpha VOC-positive samples (Ct values 23 - 29), as indicated by the higher fluorescence with the derived crRNA in samples E6 to E10 (Fig. 3e). In addition, the 69/70 deletion assay showed excellent specificity (100%), detecting all RT-qPCR positive samples and none of the RT-qPCR negative samples (Fig. 3e).

We further assessed the performance of the 69/70 assay on contrived unextracted samples. To emulate unextracted NP swab samples, we spiked in heat-inactivated viral seedstocks (reference strain or Alpha VOC) into SARS-CoV-2 negative clinical samples. Using a colorimetric lateral flow based readout, SHINEv2 correctly identified the presence or absence of the alpha VOC in the range of viral RNA concentrations (10^5^ - 10^3^ cp/μL) tested (Supplementary Fig. 15). The presence of a test band for the ancestral crRNA and lack of it for the derived crRNA confirmed the presence of the 69/70 deletion in Alpha VOC positive contrived samples with lower viral RNA concentrations (10^4^ and 10^3^ cp/μL). At higher viral RNA concentrations (10^5^ cp/μL), the presence or absence of the deletion was determined by the difference in test band intensity between the ancestral and derived crRNAs. Altogether, these data indicate that SHINEv2 can identify SARS-CoV-2 VOCs in clinical samples.

SHINEv2 is a flexible technology that can be rapidly adapted to detect new viral variants. This allowed us to develop new SHINEv2 assays for the 452 and 156 - 158 mutations present in the Delta VOC, a VOC that arose as we were preparing this manuscript for publication. We designed a set of crRNAs and RPA primers for each assay as described above, and evaluated their performance on full genome synthetic RNA standards. Both assays successfully detected the mutations in synthetic RNA targets with high sensitivity and specificity (Supplementary Fig. 16). The 156-158 assay, in particular, displayed excellent specificity. Both the ancestral and derived crRNAs detected their respective targets with high accuracy while showing no detection for the opposite target (Supplementary Fig. 16b). Next, we examined the compatibility of these assays with lyophilization and ambient-temperature sample processing. These SHINEv2 assays showed excellent specificity on full genome synthetic RNA standards, clearly distinguishing the Delta VOC from the reference strain and the other VOCs (Fig 3f). The 452 and 156-158 assays correctly identified the genotype of each variant tested, based on the differential fluorescence activity of each crRNA pair. Additionally, both SHINEv2 assays correctly identified the presence or absence of each mutation using a colorimetric lateral flow based readout (Fig. 3g and 3h). All in all, we have developed mutation-specific SHINEv2 assays which are widely deployable and, in combination, can effectively discriminate between four widely circulating SARS-CoV-2 VOCs.

### Towards at home SHINEv2 testing

SHINEv2 increases the accessibility of CRISPR-based SARS-CoV-2 diagnostics, and enables VOC identification outside of laboratory settings. However, further improvements are necessary for SHINEv2 to be used in virtually any environment. For example, the incorporation of internal controls or further simplifying the assay readout could greatly expand the use cases of SHINEv2, and enable its use in at home settings.

Nucleic acid diagnostic tests often require an internal control as part of the regulatory approval process. Therefore, we designed and developed a Cas12-based assay for human RNase P, to serve as an internal control for SHINEv2. After optimizing the concentration of RPA primers, our RNase P assay was able to detect synthetic DNA down to 100 copies/μL (Supplementary Fig. 17a and 17b). Next, we confirmed that this Cas12-based assay was compatible with the FastAmp lysis solution and with lyophilization. In this format, the Cas12-based RNase P assay detected synthetic DNA target down to 100 copies/μL using a fluorescence-based readout, which supports its potential use as an internal control for SHINEv2 (Fig. 4a). Therefore, we sought to determine the compatibility of the Cas12-based RNase P assay with the Cas13-based S gene assay and evaluate the performance of this duplex assay on synthetic nucleic acid targets. The duplex SHINEv2 assay successfully detected both targets, with an LOD of 10^3^ copies/μL for each target (Supplementary Fig. 17c). Although additional improvements will be required to achieve clinically-relevant sensitivity with the duplex SHINEv2 assay, we have taken steps towards incorporating an internal control for SHINEv2. Moreover, our results confirmed that the improvements made to SHINEv2 are applicable to other systems, including in Cas12-based assays and potentially in other CRISPR-Dx.

**Fig. 4.**
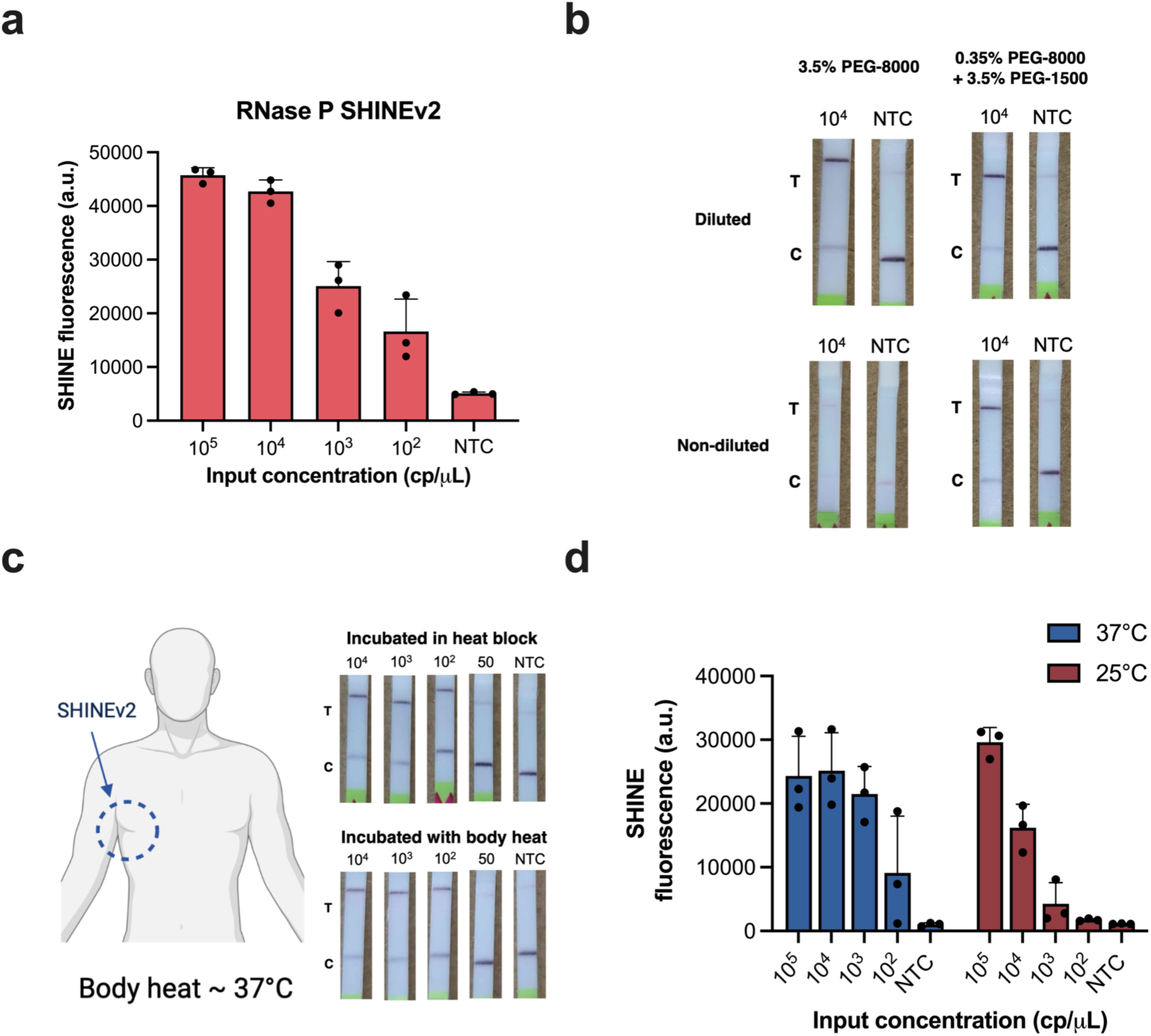
Enhancing the accessibility of SHINEv2. **a,** SHINE fluorescence of the RNase P SHINEv2 assay on synthetic DNA target after 90 minutes; NTC, no target control. **b,** Lateral flow based detection of SARS-CoV-2 RNA using SHINEv2 with different polyethylene glycol (i.e. PEG) compositions; with or without dilution after a 90 minute incubation. NTC, no target control. **c,** Lateral flow based SHINEv2 detection of SARS-CoV-2 RNA after a 90 minute incubation in a heat block or using body heat (underarm incubation). NTC, no target control. **d,** SHINE fluorescence on SARS-CoV-2 RNA after 90 minutes at 37°C or 25°C; NTC, no target control. For **d,** center = mean and error bars = s.d. for 3 technical replicates.

Moreover, the accessibility of SHINEv2 could also be enhanced by further simplifying the assay. The lateral flow based readout for CRISPR-Dx, the preferred option for point-of-need implementation, requires a buffer to be added after the detection reaction is performed, increasing the risk of sample contamination and adding a liquid-handling step for the user. Directly inserting the paper strip into the SHINEv2 reaction would significantly reduce this risk. However, undiluted SHINEv2 reactions block the flow of nanoparticles through the paper strip, resulting in a failed test (Fig. 4b). We reasoned that high molecular weight PEG – an indispensable crowding agent for RPA – could be responsible for the lack of flow. We sought to identify a combination of PEGs with different molecular weights that enables flow while retaining SHINEv2’s activity. In the absence of RNA targets, lowering the concentration and molecular weight of PEG in SHINEv2 reactions facilitated the flow of nanoparticles through the paper strip (Supplementary Fig. 18a). However, the sensitivity of SHINEv2 was markedly reduced when lowering the concentration of PEG-8000 in the reaction (Supplementary Fig. 19). Using a combination of 0.35% PEG-8000 and 3.5% PEG-1500, undiluted SHINEv2 was able to detect SARS-CoV-2 RNA without a drop in performance while retaining flow (Figure 4b and Supplementary Fig. 18b). Thus, we managed to eliminate a liquid handling step in SHINEv2 and simplify the test readout.

In addition, SHINEv2 can be run without equipment due to its minimal heating requirements. SHINE’s single 90-minute incubation step at 37°C can be easily maintained with low-cost and energy-efficient heat blocks. However, the removal of all heating elements would enable the unrestricted deployment of SHINEv2. Using body temperature, which is at approximately 37°C, could provide an alternative for fully equipment-free SHINEv2 testing^49^. Indeed, the underarm incubation of SHINEv2 was successful at detecting down to 100 cp/μL of SARS-CoV-2 RNA, and with a similar efficiency to SHINEv2 incubated in a heat block (Fig. 4c). An ideal test would perform with similar efficiency at ambient temperature as well. We examined the performance of SHINEv2 at 25°C using both a plate-based fluorescence readout and a colorimetric paper-based readout. SHINEv2 was able to detect SARS-CoV-2 RNA at 25°C with the plate reader, albeit with lower speed and sensitivity than at 37°C (Fig. 4d). We observed a larger drop in sensitivity with the lateral flow based readout (Supplementary Fig. 20). We believe the drop in sensitivity was caused by a decline in the performance of RPA at lower temperatures^50^, since Cas13 detection alone performed well at 25°C and 37°C (Supplementary Fig. 21). Collectively, these improvements increase the accessibility of SHINEv2 and facilitate its use in almost any situation.

## Discussion

Here we present SHINEv2, a widely deployable CRISPR-Dx for SARS-CoV-2 RNA detection and VOC identification from unextracted samples with a straightforward workflow. Previously developed CRISPR-Dx are not well poised for widespread deployment, as they are often too complex and/or require a cold chain and auxiliary equipment^15,20,32^. SHINEv2 addresses these limitations. Lyophilization considerably simplifies the assay and facilitates its transportation and storage. SHINEv2 can be distributed overseas without a loss in performance. Moreover, the use of an equipment-free and ambient-temperature sample lysis method further increases the user-friendliness of the assay. SHINEv2 involves as few steps from the user as antigen-capture tests, while providing a 50-fold boost in sensitivity^7,42,43^. Importantly, SHINEv2 demonstrates perfect (100%) concordance with RT-qPCR, the gold standard for SARS-CoV-2 diagnosis, in samples with RNA levels above our analytical LoD of 200 copies/μL. This level of sensitivity could enable the detection of every potentially infectious individual, including those missed by antigen-capture tests^10,46^.

SHINEv2 can accurately identify several clade-specific mutations in the Alpha, Beta, Gamma and Delta SARS-CoV-2 VOCs, and it can be rapidly adapted to respond to emerging viral variants as well as other viruses in current and future outbreaks. Thus, SHINEv2 can provide critical information to inform public health responses, and it fills a major gap in point-of-need diagnostics. At the population level, SHINEv2 could be used to prioritize testing and vaccine rollout in highly-affected communities or to select subsets of samples for further viral sequencing. SHINEv2 could also assist clinicians in selecting the right treatment (e.g. monoclonal antibody cocktails) for patients with severe COVID-19. Altogether, we believe that SHINEv2 will be especially valuable for community surveillance testing, as it combines user-friendly and equipment-free preparation methods with sufficient sensitivity and the capacity to discriminate VOCs.

Further advances will be required for CRISPR-based diagnostic testing to take place in any location, including at-home settings. Ideally, such a test would involve a few simple, ambient-temperature steps and provide a fast and accurate visual readout without the need for specialized equipment. To our knowledge, current nucleic acid diagnostics are not capable of meeting all these requirements simultaneously. The combination of sample processing, nucleic acid amplification and CRISPR-based detection into a single, ambient-temperature reaction could remove liquid handling steps. The incorporation of solution-based, colorimetric readouts could further simplify the assay and reduce the risk of contamination^51^. Moreover, with SHINEv2, we have taken steps to eliminate all equipment needs, although additional advances will be required to boost SHINEv2’s performance at ambient temperature. The speed and sensitivity of ambient-temperature CRISPR-Dx could be enhanced through the addition of auxiliary proteins or using alternative isothermal amplification methods^18,21,52^. Collectively, these improvements could greatly enhance the accessibility of SHINEv2 and provide a critical tool in the fight against current and future outbreaks. By reducing assay complexity and simplifying test distribution, without sacrificing sensitivity or specificity, we have taken steps towards the development of such a diagnostic tool.

## Methods

### Clinical samples and ethics statement

This study received a non-human subjects research determination from the Broad Institute Office of Research Subject Protections (NHSR-4318). To conduct this research, de-identified nasopharyngeal swab patient samples were purchased from Boca Biolistics (USA). Additional de-identified clinical samples were obtained from studies evaluated and approved at the Center for Disease Control and Prevention (CDC). This activity was reviewed by CDC and was conducted consistent with applicable federal law and CDC policy (See e.g., 45 C.F.R. part 46; 21 definition of research as defined in and was conducted consistent with C.F.R. part 56; 42 U.S.C. 241(d), 46.102(l) but did not involve human applicable federal law and CDC U.S.C. 552a, 44 U.S.C. 3501 et seq.)

### SARS-CoV-2 assay design and synthetic template information

RPA primers were designed using ADAPT^36^. The S gene, which contains SNPs of interest in several SARS-CoV-2 VOCs, was extracted from an alignment of 308,315 SARS-CoV-2 genomes downloaded from GISAID in January 2021^53,54^. ADAPT was then run on the S gene alignment with the following parameters: 35-65% GC content, max. 1 mismatch between the primer and target, full coverage in >98% of the genomes and overlap allowed between amplicons but not primers. Forward RPA primers were ordered with a T7 promoter sequence (5’-GAAATTAATACGACTCACTATAGGG-3’) appended upstream; and both forward and reverse RPA primers were purchased from Integrated DNA Technologies (IDT) as single-stranded DNA oligos.

The CRISPR RNAs (crRNAs) for SNP discrimination were designed using a generative sequence design algorithm^55^. This approach uses ADAPT’s predictive model to predict the activity of candidate crRNA sequences against on-target and off-target sequences^36^. These predictions of candidate crRNA activity guide the generative algorithm’s optimization process, in which it seeks to design crRNA probes that have maximal predicted on-target activity and minimal predicted off-target activity. These crRNAs were purchased from IDT as Alt-R CRISPR guide RNAs.

Synthetic DNA targets with an upstream T7 promoter sequence were ordered as double-stranded DNA (dsDNA) gene fragments from IDT, and were *in vitro* transcribed to generate synthetic RNA targets. *In vitro* transcription was conducted as previously described^23^. In brief, the dsDNA template was transcribed using the HiScribe T7 High Yield RNA Synthesis Kit (New England Biolabs (NEB)) for 2 hours at 37 °C. Transcribed RNA was then treated with RNase-free DNase I (QIAGEN) according to the manufacturer’s instructions. Finally, DNase-treated RNA was purified using RNAClean SPRI XP beads using a 10:3:5 mixture of RNA solution, isopropanol and ethanol.

Sequence information for the synthetic targets, RPA primers, Cas13-crRNA and reporters is listed in Supplementary Table 2.

### Full-genome synthetic RNA controls and viral seedstocks

Synthetic SARS-CoV-2 RNA controls (controls 2, 14, 16, 17 and 23) were purchased from Twist Biosciences and are referred to as full genome synthetic RNA standards.

Heat-inactivated viral stocks (2019-nCoV/USA-WA1/2020 as reference strain and USA/CA_CDC_5574/2020 as Alpha VOC) were purchased from ATCC.

### Analysis of crRNA activity with Cas13a

Initial measurements of crRNA activity were obtained under simplified SHINE conditions. That is, crRNA activity was measured using 1X optimized reaction buffer (20 mM HEPES pH 8.0 with 60 mM KCl and 3.5% PEG-8000), 45 nM *Lwa*Cas13a protein, 125 nM polyU [*i.e.*, 6 uracils (6U) in length] quenched FAM reporter, 1 U/µL murine RNase inhibitor, 22.5 nM crRNA and 14mM magnesium acetate. Target RNA was added to each reaction at a ratio of 1:20 and fluorescence was measured on a Biotek Cytation 5 plate reader (excitation: 485 nm, emission: 520 nm) at 37 °C every 5 minutes for up to 3 hours.

### Nuclease inactivation with FastAmp Lysis reagent

A concentrated (5X) version of FastAmp Viral and Cell solution B for Covid-19 Testing was purchased directly from Intact Genomics. This solution, referred to as FastAmp Lysis reagent, was used at a final concentration of 1X. For nuclease inactivation experiments, 10% healthy nasal fluid (Lee BioSolutions) in universal transport medium (UTM) was mixed with 1X lysis reagent and either 1%, 2% or 5% (v/v) murine RNase inhibitor (NEB). HUDSON treatment was used as a control for successful RNase inactivation and performed as previously described^23^. In short, 100 mM TCEP (Thermo Fisher Scientific), 1 mM EDTA (Thermo Fisher Scientific) and 0.8 U/µL murine RNase inhibitor were added to the nasal fluid/UTM mix and incubated at 40 °C for 5 minutes. PBS was used instead of HUDSON reagents as a no-treatment control. All treated products were then mixed 1:1 with 400 mM RNaseAlertv2 substrate (Thermo Fisher Scientific) in nuclease-free water and incubated at 25°C, while measuring the fluorescence (excitation: 485 nm, emission: 520 nm) every 5 minutes for up to 30 minutes using a Biotek Cytation 5 plate reader.

### Viral inactivation with Intact Genomics Lysis buffer

2019-nCoV/USA-WA1-A12/2020 isolate of SARS-CoV-2 was obtained from the US Centers for Disease Control and Prevention (CDC). At the Integrated Research Facility (IRF) - Frederick, the virus was passaged by inoculating grivet kidney epithelial Vero cells (ATCC #CCL-81) at a multiplicity of infection (MOI) of 0.01 under high containment (BSL-3) conditions. Infected cells were incubated for 48 or 72 hours in Dulbecco’s Modified Eagle Medium with 4.5g/L D-glucose, L-glutamine, and 110 mg/L sodium pyruvate (DMEM, Gibco) containing 2% heat-inactivated fetal bovine serum (SAFC Biosciences) in a humidified atmosphere at 37°C with 5% CO_2_. The resulting viral seedstock was harvested and quantified by plaque assay using Vero E6 cells (ATCC #CRL-1586) with a 2.5% Avicel overlay and stained after 48 hours with a 0.2% crystal violet stain.

For testing the viral inactivation capacity of FastAmp Lysis reagent, the viral stock (10^7^ PFU/mL) was mixed with 1X lysis buffer supplemented with 5% (v/v) murine RNAse inhibitor and incubated at ambient temperature for either A) 5 minutes, B) 20 minutes or C) 20 minutes followed by 10 minutes at 65°C. A no-treatment (PBS) control was also included. Treated viral stocks were then cleaned and quantified by plaque assay using Vero E6 cells (ATCC #CRL-1586) with a 2.5% Avicel overlay and stained after 48 hours with a 0.2% crystal violet stain.

### Single-step SARS-CoV-2 SHINE reactions

RPA primer and crRNA optimizations were performed using the following SHINE conditions: 1X original SHINE buffer (20 mM HEPES pH 8.0 with 60 mM KCl and 3.5% PEG-8000), 45 nM *Lwa*Cas13a protein resuspended in 1X SB (such that the resuspended protein is at 2.26 µM), 125 nM polyU quenched FAM reporter, 2 mM of each rNTP, 1 U/µL murine RNase inhibitor, 1 U/µL NextGen T7 RNA polymerase, 0.1 U/µL RNase H (NEB), 2 U/µL Invitrogen SuperScript IV (SSIV) reverse transcriptase (Thermo Fisher Scientific), an assay specific concentration of forward and reverse RPA primers (detailed below), and 22.5 nM crRNA. Once the master mix is created, it is used to resuspend the TwistAmp Basic Kit RPA pellets (1 lyophilized pellet per 102 µL final master mix volume). After that, magnesium acetate (as the only magnesium cofactor) is added at a final concentration of 14 mM to generate the final master mix. Finally, a synthetic RNA target or a sample was added to the complete master mix at a ratio of 1:19. Fluorescence kinetics were measured on a Biotek Cytation 5 plate reader (excitation: 485 nm, emission: 520 nm) at 37 °C every 5 minutes for up to 3 hours.

For lateral flow readout, the quenched FAM reporter was exchanged for a biotinylated FAM reporter at a final concentration of 1 µM. After incubation at 37°C for 90 minutes, the SHINE reaction was diluted 1:4 in 80μL HybriDetect Assay Buffer (Milenia Biotec), and a HybriDetect 1 lateral flow strip was added. Test images were collected 5 minutes after the addition of the strip with a smartphone camera.

RPA primer concentrations varied between assays, but the ratio of forward to reverse primers was always kept to 1:1. For each assay, the concentration of forward and reverse RPA primer was the following: 120nM for ORF1a and 417 assays; 160nM for 452 assay and 180nM for S gene, 69/70 deletion and 156-158 assays.

### SHINE tolerance to samples inactivated with lysis solution

Contrived nasal samples (10% (v/v) in UTM) were inactivated with lysis solution (FastAmp lysis reagent with 5% RNase inhibitor) for 5 minutes at ambient temperature. SHINE reactions were generated as described above, except a variable amount of water was replaced with inactivated contrived nasal samples. Synthetic RNA target (with the concentration specified in the figure) was added to the modified SHINE reactions at a ratio of 1:19. Plate-based fluorescence detection was performed as described above. In these reactions, the inactivated sample volume comprised 0% - 40% of the total volume.

### Lyophilization optimization for SHINE

The original SHINE buffer was optimized to retain SHINE’s activity after lyophilization. For lyophilization, we generated the master mix as described above, except the 1X SHINE buffer was substituted for 1X lyophilization buffer (composition described below) and magnesium acetate was omitted. The mastermix was aliquoted, flash frozen in liquid nitrogen and lyophilized overnight. The lyophilization buffers tested had the following composition: 1) SHINE (20 mM HEPES pH 8.0 with 60 mM KCl and 3.5% PEG-8000); 2) SHINE without PEG and KCl (20 mM HEPES pH 8.0), 3) SHINE with sucrose and mannitol (20 mM HEPES pH 8.0 with 60 mM KCl, 3.5% PEG-8000, 5% (w/v) sucrose and 150mM mannitol); 4) LYO (20 mM HEPES pH 8.0 with 5% w/v sucrose and 150mM mannitol). Lyophilized pellets were resuspended with a buffer composed of magnesium acetate (14 mM final concentration) and whichever reagent was left out of the original SHINE buffer (e.g. 60 mM KCl, 3.5% PEG-8000 and 14 mM magnesium acetate for the LYO resuspension buffer (RB)). After resuspension, synthetic RNA target was added to the resuspended SHINE reactions at a ratio of 1:19. Plate-based fluorescence detection or lateral flow based detection were performed as described above.

Lyophilized SHINE pellets for stability testing were prepared using the LYO buffer as described above. SHINE pellets were stored at room temperature, 4°C or -20°C for 4,7,14,21 or 28 prior to resuspension and testing. For SHINEv2 testing in Nigeria, SHINE pellets were lyophilized in the United States, shipped in dry ice to Nigeria and then resuspended and tested after a total of 6 weeks in storage.

### Side-by-side clinical sample testing

Clinical samples were thawed on ice. SHINEv2, BinaxNow and CareStart testing as well as RNA extractions and RT-qPCR were performed side-by-side, according to the manufacturer’s instructions (see details below). 96 nasopharyngeal swab samples were tested in batches of 24 samples.

Information on clinical samples, including C_t_ values, can be found in Supplementary Table 1. Clinical sample test results in Supplementary Fig. 8 - 10.

### Clinical sample extractions and RT-qPCR

Clinical sample extractions and RT-qPCR were performed according to the CDC’s recommended protocol^41^. Nucleic acid extractions for samples S1 - S96 were performed using the QIAamp Viral RNA Mini Kit (Qiagen). The starting volume for extraction was 100 μL and extracted nucleic acid was eluted into 100 μL of nuclease-free water. Extracted RNA was used immediately or stored at -80C. Extracted RNA was tested for the presence of SARS-CoV-2 RNA using the CDC’s SARS-CoV-2 RT-qPCR assay (2019-nCoV CDC EUA Kit, IDT). RT-qPCR cycling conditions were as follows: hold at 25°C for 2 min, reverse transcription at 50 °C for 15 min, polymerase activation at 95 °C for 2 min and 45 cycles with a denaturing step at 95 °C for 3 s followed by annealing and elongation steps at 55 °C for 30 s. RT-qPCR was run on a QuantStudio 6 (Applied Biosystems) and data were analyzed using the Standard Curve (SC) module of the Applied Biosystems Analysis Software.

### SHINEv2 testing - clinical samples

SHINEv2 reactions were prepared the day before testing and consist of 1X optimized lyophilization buffer (20 mM HEPES pH 8.0 with 5% w/v sucrose and 150mM mannitol), 45 nM *Lwa*Cas13a protein resuspended in 1X SB (such that the resuspended protein is at 2.26 µM), 1 µM lateral flow biotinylated FAM reporter, 2 mM of each rNTP, 1 U/µL murine RNase inhibitor, 1 U/µL NextGen T7 RNA polymerase, 0.1 U/µL RNase H (NEB), 2 U/µL Invitrogen SuperScript IV (SSIV) reverse transcriptase (Thermo Fisher Scientific),180nM forward and reverse RPA primers and 22.5 nM crRNA. Once the master mix was created, it was used to resuspend the TwistAmp Basic Kit lyophilized reaction components (1 lyophilized pellet per 102 µL final master mix volume). The mastermix was aliquoted into individual reactions (20μL final), flash frozen in liquid nitrogen and lyophilized overnight.

The following day, clinical samples were mixed with lysis reagent (diluted to 1X) with 5% RNase inhibitor and incubated at ambient temperature for 5 minutes. In the meantime, the lyophilized SHINEv2 pellets were resuspended with Resuspension Buffer to 1X (60 mM KCl, 3.5% PEG-8000 and 14mM magnesium acetate). Inactivated clinical samples were added to the resuspended SHINEv2 reactions at a ratio of 1:9. After incubation at 37C for 90 minutes, the SHINEv2 reaction was diluted in 80μL HybriDetect Assay Buffer, and a HybriDetect 1 lateral flow strip was added. Test images were collected 5 minutes after the addition of the strip. Tests were considered positive when the intensity of the test band was higher than that of the negative controls.

### BinaxNow COVID - 19 Antigen self-test - clinical samples

BinaxNow COVID-19 Antigen self-tests (Abbott) were purchased from Walmart. Since we were not able to procure dry swabs, the test was performed according to the manufacturers’ protocol for determining the analytical sensitivity with a liquid input (Instructions For Use submitted for FDA EUA)^56^. Briefly, 6 drops of BinaxNow liquid were added to the top hole of each test card. 20μL of clinical samples were absorbed onto a clinical swab. The swab was then inserted into the bottom hole of the test card and turned 3 times. Without removing the swab, the test card was closed and sealed. The test was incubated at room temperature for at least 15 minutes before imaging with a smartphone camera. Any faint colored lines in the test region were considered as positive.

### CareStart COVID-19 Antigen testing - clinical samples

CareStart COVID-19 Antigen tests (Access Bio) were purchased from Medek Health. The test was performed according to the manufacturer instructions for use^57^. Briefly, clinical samples were mixed 1:1 with the extraction buffer. 3 drops of that mix were added onto the sample well in a test cartridge and incubated at ambient temperature for at least 10 minutes before imaging with a smartphone camera. As suggested by the manufacturer, any faint colored line in the test region was considered as positive.

### Contrived samples testing (VOCs)

Contrived samples were created by diluting a known concentration of heat-inactivated viral seedstocks (reference strain or Alpha VOC) 1:9 in UTM. Contrived samples were then inactivated with lysis solution for 5 minutes at ambient temperature. Lyophilized SHINEv2 pellets were prepared and resuspended as described above. Inactivated samples were added to the resuspended SHINEv2 reactions at a ratio of 1:9. After incubation at 37°C for 90 minutes, the SHINEv2 reaction was diluted in 80μL HybriDetect Assay Buffer, and a Milenia HybriDetect 1 lateral flow strip was added. Test images were collected 5 minutes after the addition of the strip.

### Extracted clinical samples testing (VOCs)

Clinical sample extractions and RT-qPCR were performed according to the CDC’s recommended protocol^41^. Nucleic acid extraction for samples E6 - E10 were performed with the MagnaPure 96 instrument using the DNA and viral SV kit (Roche). The starting volume for extraction was 100 μL and extracted nucleic acid was eluted into 100 μL of nuclease-free water. Extracted RNA was used immediately or stored at -80C. Extracted RNA was tested for the presence of SARS-CoV-2 RNA using the CDC’s SARS-CoV-2 RT-qPCR assay (2019-nCoV CDC EUA Kit, IDT). RT-qPCR cycling conditions were as follows: hold at 25°C for 2 min, reverse transcription at 50 °C for 15 min, polymerase activation at 95 °C for 2 min and 45 cycles with a denaturing step at 95 °C for 3 s followed by annealing and elongation steps at 55 °C for 30 s. RT-qPCR was run on a QuantStudio 6 (Applied Biosystems) and data were analyzed using the Standard Curve (SC) module of the Applied Biosystems Analysis Software.

RNA extracted from clinical samples was added to resuspended SHINEv2 reactions at a ratio of 1:9, as described above. After incubation at 37°C for 90 minutes, the SHINEv2 reaction was diluted in 80μL HybriDetect Assay Buffer, and a HybriDetect 1 lateral flow strip was added. Test images were collected 5 minutes after the addition of the strip.

### Cas12-based RNase P assay

Cas12-based SHINEv2 reactions were prepared as follows: 1X original SHINE buffer or LYO buffer, 20 nM *Lba*Cas12a, 250nM polyC [*i.e.*, 5 cytosines (5C) in length] quenched HEX reporter, 1 U/µL murine RNase inhibitor, 90nM of forward and reverse RPA primers, and 22.5 nM Cas12a crRNA were combined to create a master mix. The master mix was used to resuspend the TwistAmp Basic Kit RPA pellets (1 lyophilized pellet per 102 µL final master mix volume). For lyophilization, the magnesium acetate was omitted, and individual reactions were aliquoted, flash frozen and lyophilized overnight. Otherwise, magnesium acetate (as the only magnesium cofactor) was added at a final concentration of 14 mM. Lyophilized Cas12 reactions were reactivated with Resuspension Buffer, as described above. Finally, synthetic DNA target was added to the complete master mix at a ratio of 1:19. Fluorescence kinetics were measured on a Biotek Cytation 5 plate reader (excitation: 485 nm, emission: 520 nm) at 37 °C every 5 minutes for up to 3 hours. For lateral flow readout, the quenched HEX reporter was exchanged for a 5C biotinylated FAM reporter at a final concentration of 1 µM. After incubation at 37°C for 90 minutes, the Cas12-based reaction was diluted in 80μL HybriDetect Assay Buffer, and a HybriDetect 1 lateral flow strip was added. Test images were collected 5 minutes after the addition of the strip.

### Duplex SHINEv2

Duplex SARS-CoV-2 and RNase P SHINEv2 reactions were prepared as follows: 1X original SHINE buffer or 1X LYO buffer, 45 nM *Lwa*Cas13a, 20nM *Lba*Cas12a, 125 nM polyU quenched FAM reporter, 250nM polyC quenched HEX reporter, 2 mM of each rNTP, 1 U/µL murine RNase inhibitor, 1 U/µL NextGen T7 RNA polymerase, 0.1 U/µL RNase H, 2 U/µL Invitrogen SuperScript IV (SSIV) reverse transcriptase, 90nM S gene RPA primers, 90nM RNase P RPA primers, 22.5 nM Cas12a crRNA and 22.5 nM Cas13a crRNA were combined to create a master mix. The mastermix was then used to resuspend the TwistAmp Basic Kit RPA pellets (1 lyophilized pellet per 102 µL final master mix volume). After that, pre- and post-lyophilization reactions were performed as previously described.

### PEG experiments

To evaluate the effect of PEG composition and concentration on flow through the lateral flow strip, SHINEv2 reactions were prepared as described above, except the 3.5% PEG-8000 present in the original SHINE buffer was exchanged for 0 - 10% PEG-8000, PEG-1500 or a combination of the two. Without adding synthetic targets and with or without diluting in HybriDetect Assay Buffer, a Milenia HybriDetect 1 lateral flow strip was added to these modified reactions. Test images were collected 5 minutes after the addition of the strip.

To evaluate the performance of SHINEv2 with modified PEG composition, SHINEv2 reactions were performed as described above, except the composition of the original SHINE and resuspension buffers was modified to be SHINE (20 mM HEPES pH 8.0 with 60 mM KCl, 0.35% PEG-8000 and 3.5% PEG-1500) and resuspension buffer (60 mM KCl, 0.35% PEG-8000, 3.5% PEG-1500 and 14mM magnesium acetate).

### Body heat experiments

Lyophilized SHINEv2 pellets were prepared and resuspended as described above. Full genome synthetic RNA standards were added to individual SHINEv2 reactions and incubated using body heat or a heat block for 90 minutes. For body heat experiments, SHINEv2 reactions were closed, inserted in a ziploc bag, taped to the underarm of one of the researchers and incubated for 90 minutes. The reactions were then diluted in 80μL HybriDetect Assay Buffer, and a HybriDetect 1 lateral flow strip was added to each reaction. Test images were collected 5 minutes after the addition of the strip.

### Room temperature experiments

Lyophilized SHINEv2 pellets were prepared and later resuspended as described above. Contrived nasal samples were inactivated with lysis solution and added to the resuspended SHINEv2 reactions at a ratio of 1:9. Fluorescence kinetics were measured on a Biotek Cytation 5 plate reader (excitation: 485 nm, emission: 520 nm) every 5 minutes for up to 3 hours, at either 37 °C or 25 °C.

### Data analysis and schematic generation

SHINE fluorescence values are reported as background-subtracted fluorescence, with the fluorescence value collected before reaction progression (usually t = 10 minutes) subtracted from the final fluorescence value (90 minutes, unless otherwise indicated). Since the SHINEv2 VOC data was collected using different plate-readers, SHINE fluorescence in Fig. 3 was normalized. For normalization, the maximal fluorescence value in an experiment is set to 1 and the fluorescence values from that same experiment are set as ratios of the max. fluorescence value.

Schematics shown in Fig. 1a, 1d, 2a, 3a, 4c and Supplementary Fig. 5 were created using www.biorender.com. Data panels were primarily generated using Prism 8 (GraphPad).

## Data Availability

All data produced in the present work are contained in the manuscript.

## Acknowledgements

We would like to thank the TIDE group at the Broad Institute for providing additional laboratory space to perform the work; all the researchers and laboratories who generously made SARS-CoV-2 sequencing data publicly available, to inform the design of our assay; H. Metsky, for his contributions to the development of ADAPT, which guided the assay design; the Sabeti laboratory, notably S. Siddiqui, H. Metsky, E. Normandin for their thoughtful discussions and reading of the manuscript. Funding was provided by DARPA D18AC00006. This work is made possible by support from Flu Lab and a cohort of generous donors through TED’s Audacious Project, including the ELMA Foundation, MacKenzie Scott, the Skoll Foundation, and Open Philanthropy. J.A.-S. is supported by a fellowship from “la Caixa” Foundation (ID 100010434, code LCF/BQ/AA18/11680098). C.M. is supported by start-up funds from Princeton University. For L.W., this work was funded under Agreement No. HSHQDC-15-C-00064 awarded to Battelle National Biodefense Institute (BNBI) by the Department of Homeland Security (DHS) Science and Technology (S&T) Directorate for the management and operation of the National Biodefense Analysis and Countermeasures Center (NBACC), a Federally Funded Research and Development Center.

The views, opinions, conclusions, and/or findings expressed should not be interpreted as representing the official views or policies, either expressed or implied, of the Department of Defense, US government, National Institute of General Medical Sciences, DHS, or the National Institutes of Health. The DHS does not endorse any products or commercial services mentioned in this presentation. In no event shall the DHS, BNBI or NBACC have any responsibility or liability for any use, misuse, inability to use, or reliance upon the information contained herein. In addition, no warranty of fitness for a particular purpose, merchantability, accuracy or adequacy is provided regarding the contents of this document. Notice: This manuscript has been authored by Battelle National Biodefense Institute, LLC under Contract No. HSHQDC-15-C-00064 with the U.S. Department of Homeland Security. The United States Government retains and the publisher, by accepting the article for publication, acknowledges that the United States Government retains a non-exclusive, paid up, irrevocable, world-wide license to publish or reproduce the published form of this manuscript, or allow others to do so, for United States Government purposes.

## Contributions

J.A.-S. and C.A.F. conceived this study under the guidance of P.C.S. and C.M. J.A.-S., A.B, C.K.B, C.A.F., M.E.G., T.-S.F.K.-T. and G.K. performed initial experiments to improve the SHINE technology. G.K. and R.G. performed viral inactivation experiments, under the guidance of L.E.H. J.A-S., A.B., C.K.B. and M.E.G performed experiments and data analysis related to the S gene assay and the development of SHINEv2. J.N.U, P.E.E and O.G.J. performed SHINEv2 experiments in Nigeria, under the supervision of C.T.H. J.A.-S., A.B., N.L.W, P.P.P. and S.M. designed and/or tested SHINEv2 assays for SARS-CoV-2 variant detection. J.S.L. provided assistance in patient sample collection. J.A.-S. and A.B performed and analyzed experiments with patient samples. Y.B.Z. designed and validated Cas12-based SHINEv2 assays. L.E.H., C.T.H. and J.J. provided critical insights on protocols, the experimental results, and the work overall. J.A.-S. wrote the paper with guidance from P.C.S. and C.M. P.C.S and C.M. jointly supervised the project. All authors reviewed the manuscript.

## Supplementary material

**Supplementary Fig. 1.**
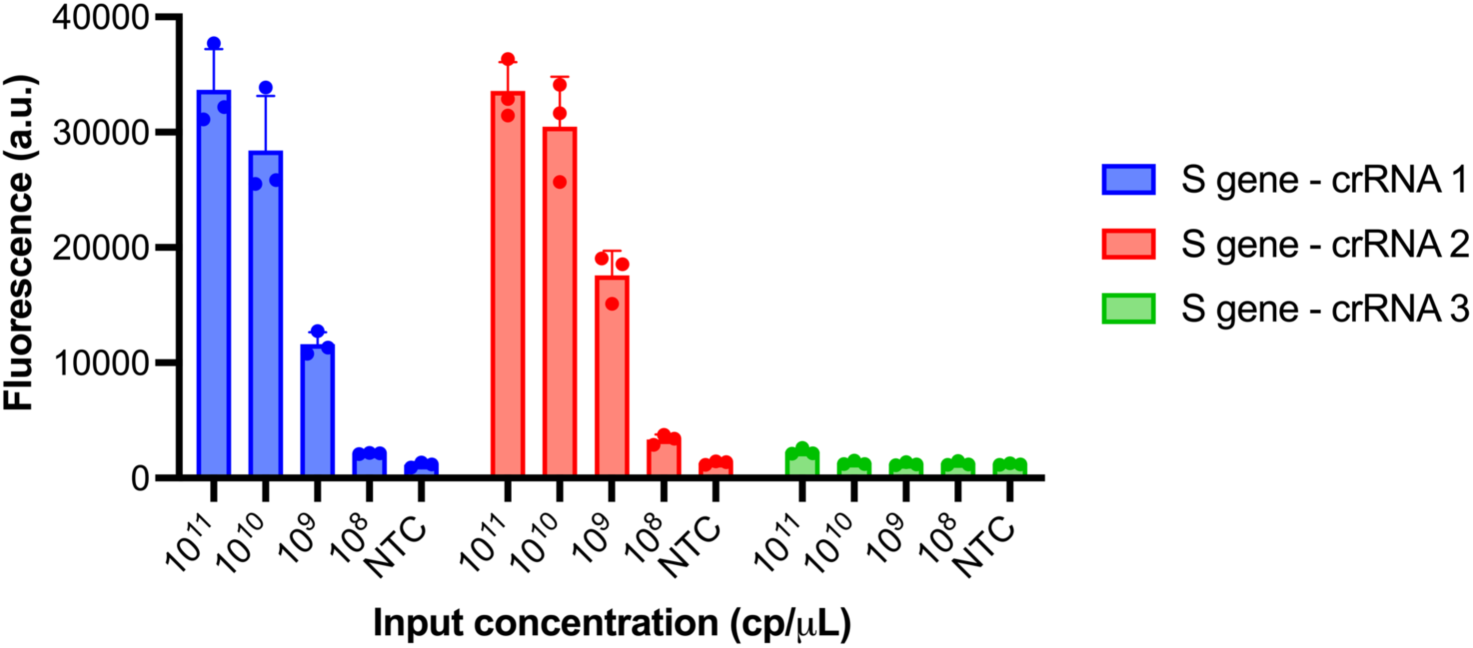
Selection of most active S gene crRNA. Cas13a detection of synthetic RNA target (S gene) using different crRNAs. NTC, no target control. Centre = mean and error bar = standard deviation (s.d.) for 3 technical replicates.

**Supplementary Fig. 2.**
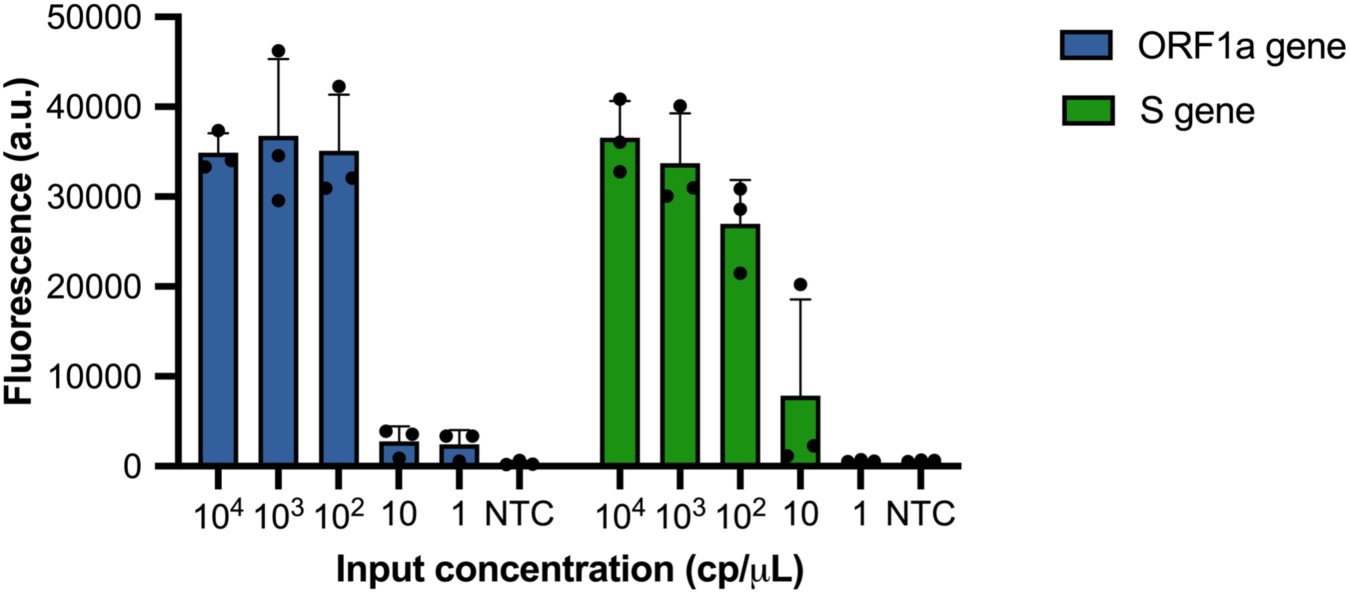
SHINE assay comparison. Background-subtracted fluorescence of the ORF1a and S gene SHINE assays using synthetic SARS-CoV-2 RNA after 90 minutes. NTC, no target control. Center = mean and error bars = s.d. for 3 technical replicates.

**Supplementary Fig. 3.**
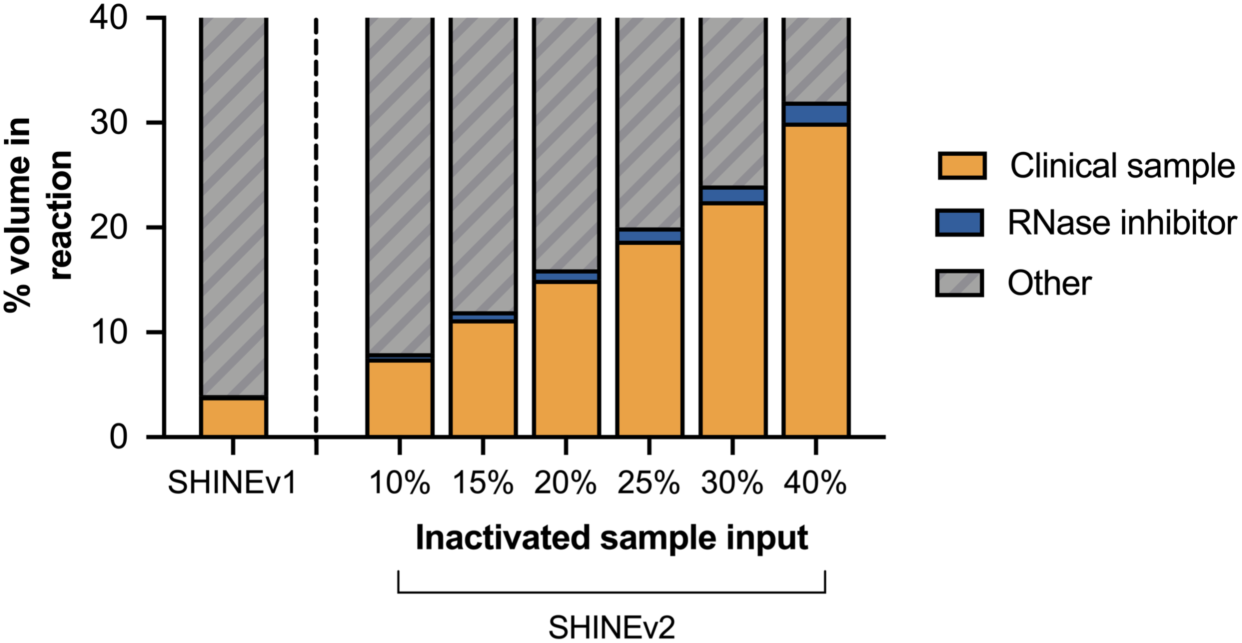
Inactivated sample input is increased in SHINEv2. Volume (%) of clinical sample and RNase inhibitors in the final reaction for SHINEv1 and SHINEv2 as a function of inactivated sample input.

**Supplementary Fig. 4.**
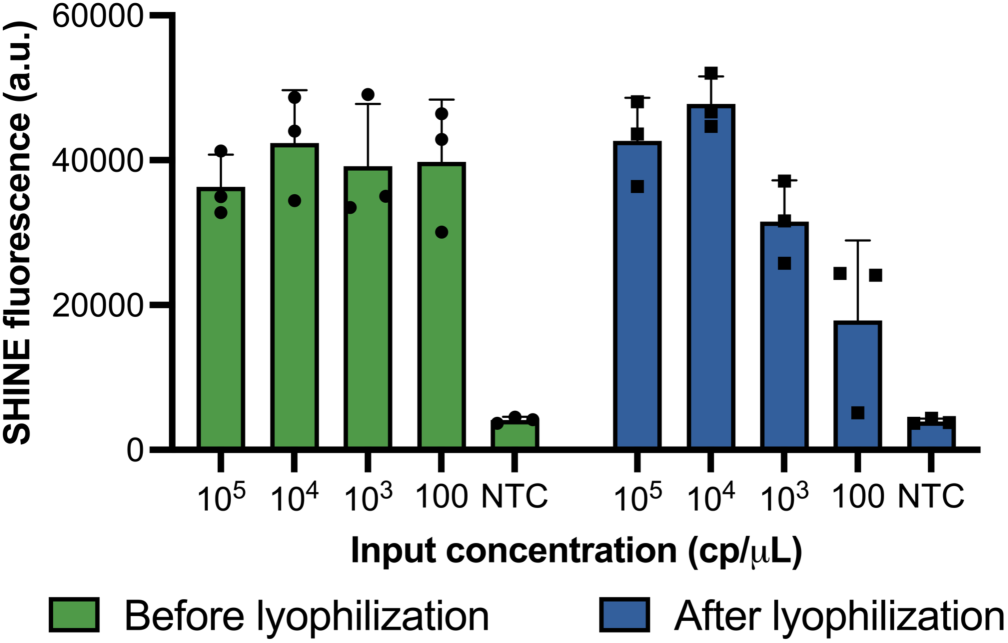
The activity of SHINE is preserved after lyophilization. SHINE fluorescence on synthetic RNA target before and after lyophilization. Fluorescence measured after 90 minutes. NTC, no target control. Center = mean and error bars = s.d. for 3 technical replicates.

**Supplementary Fig. 5.**
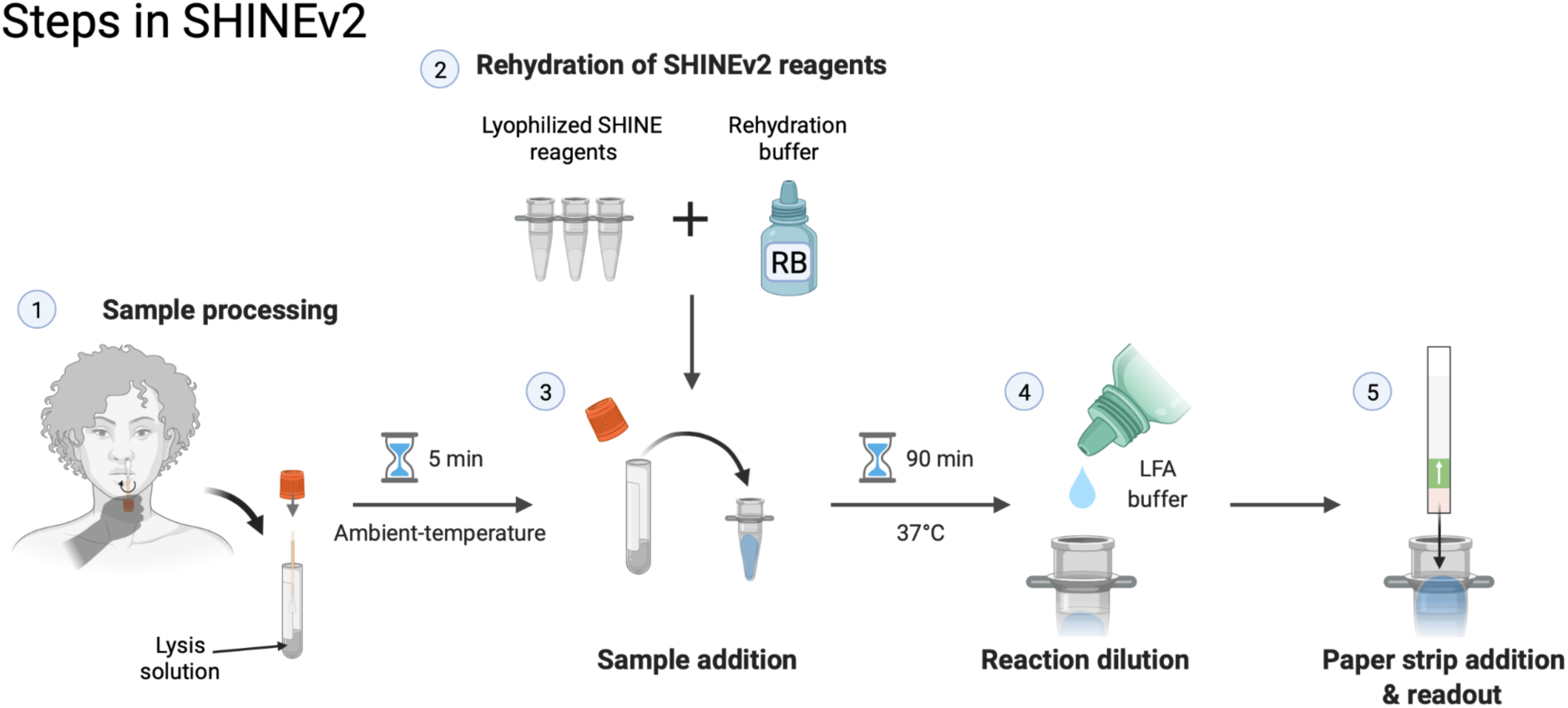
SHINEv2 requires 5 user manipulations. Schematic of the SHINEv2 workflow, including sample inactivation, rehydration of pellets, sample addition, reaction dilution after incubation and result readout.

**Supplementary Fig. 6.**
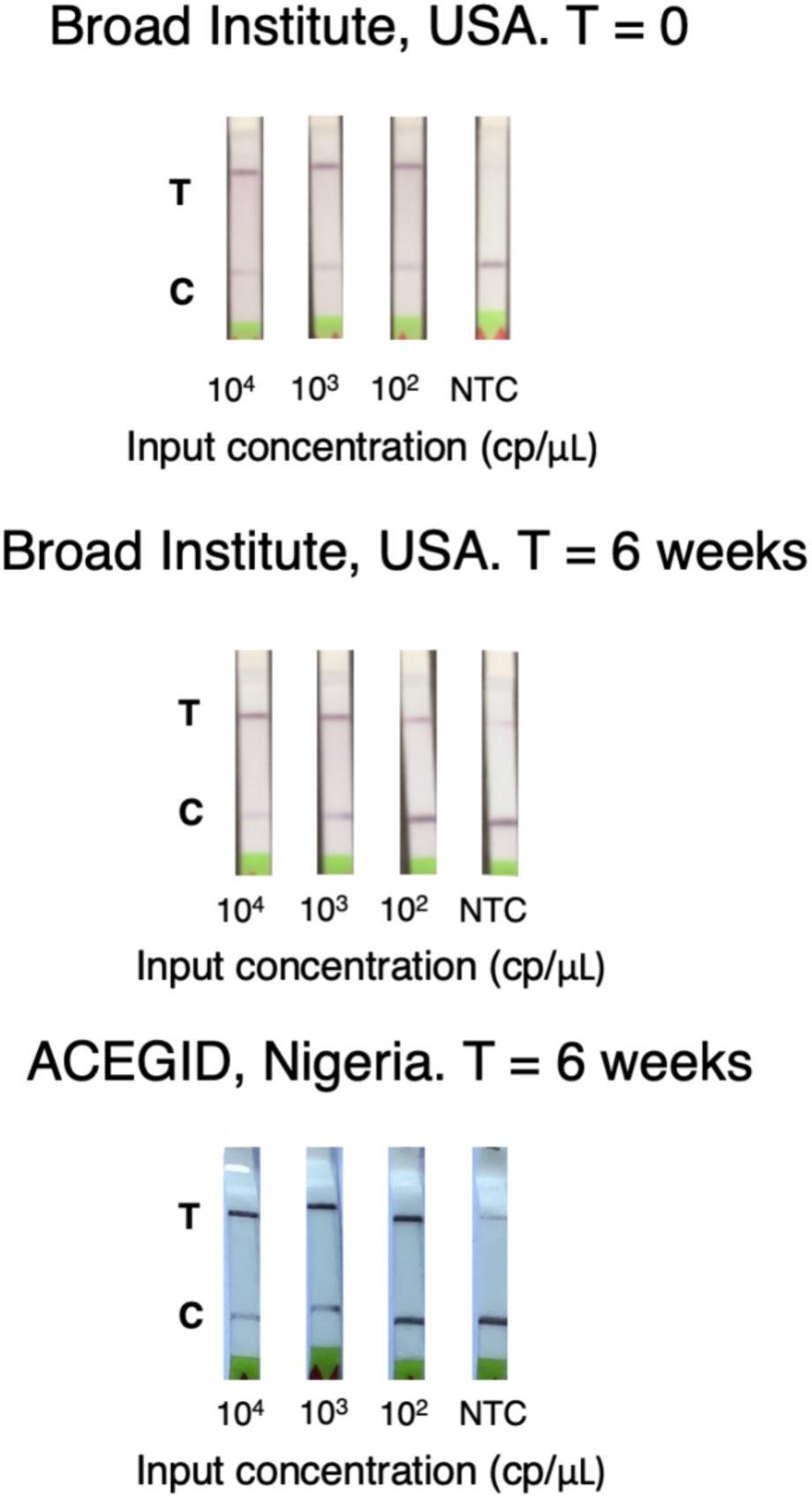
SHINEv2 can be transported overseas and is stable for at least 6 weeks. Lateral-flow detection of full-genome synthetic RNA standards in lysis solution-treated UTM using SHINEv2, before and after transportation. Incubated for 90 minutes. C = control band; T = test band; NTC = no target control.

**Supplementary Fig. 7.**
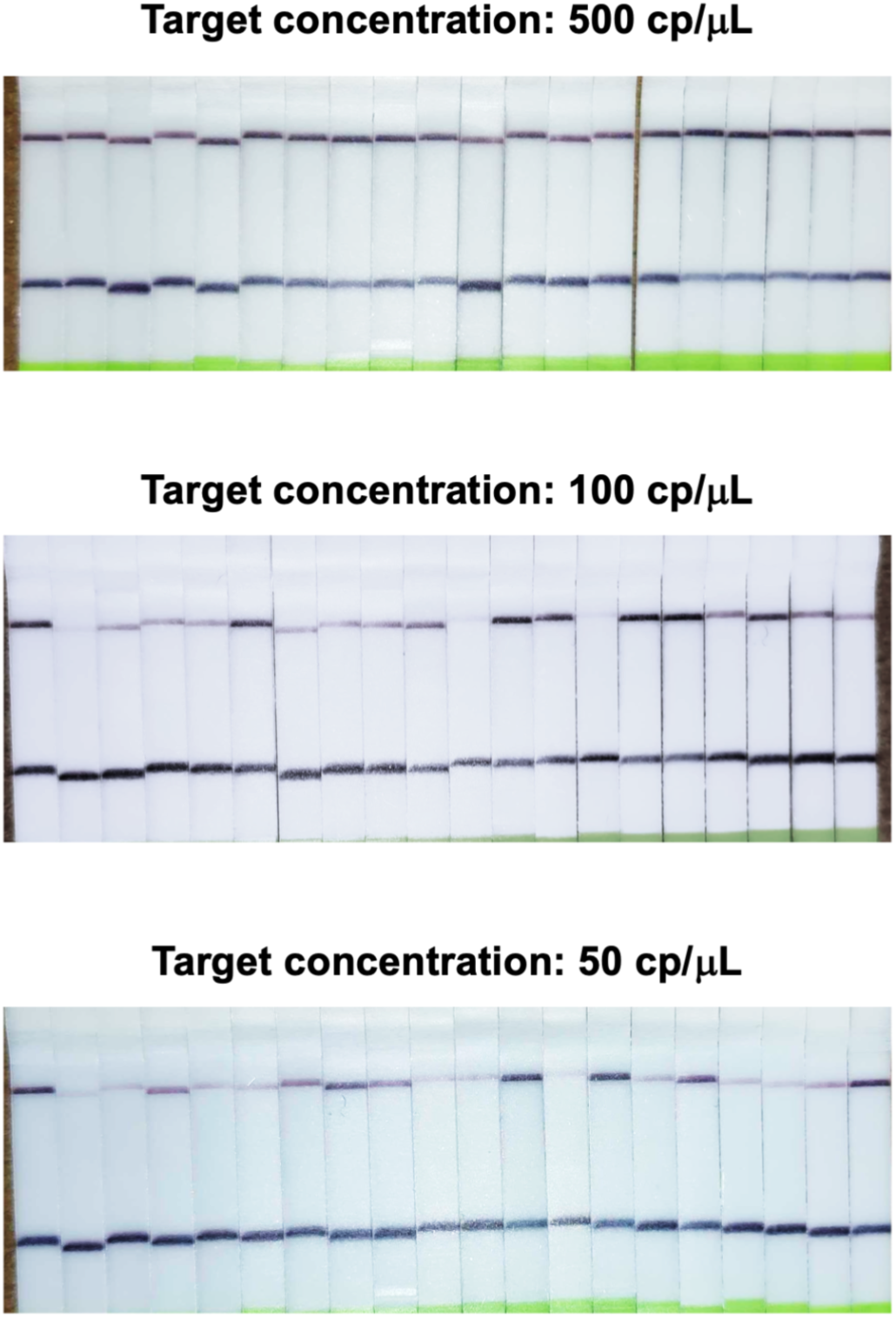
SHINEv2 has an analytical sensitivity of 200cp/μL. Determination of analytical limit of detection with 20 replicates of SHINEv2 at different concentrations of SARS-CoV-2 RNA from lysis solution-treated contrived samples. Incubated for 90 minutes. Results for 200cp/μL sample input shown in Fig. 1i.

**Supplementary Fig. 8.**
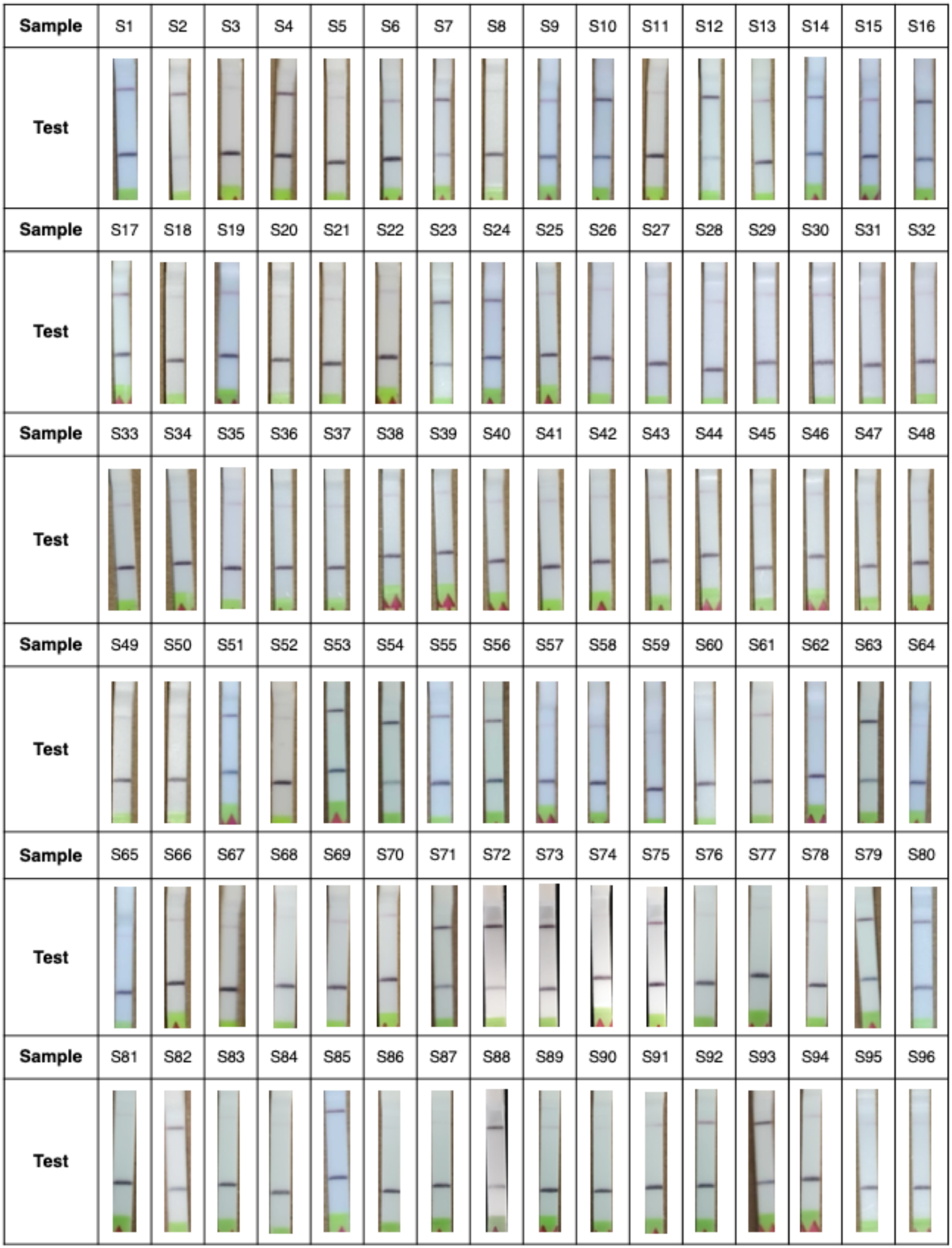
SHINEv2 results on clinical samples. Images of positive and negative test results with SHINEv2 on unextracted nasopharyngeal (NP) swab samples. Samples were incubated for 90 minutes at 37°C.

**Supplementary Fig. 9.**
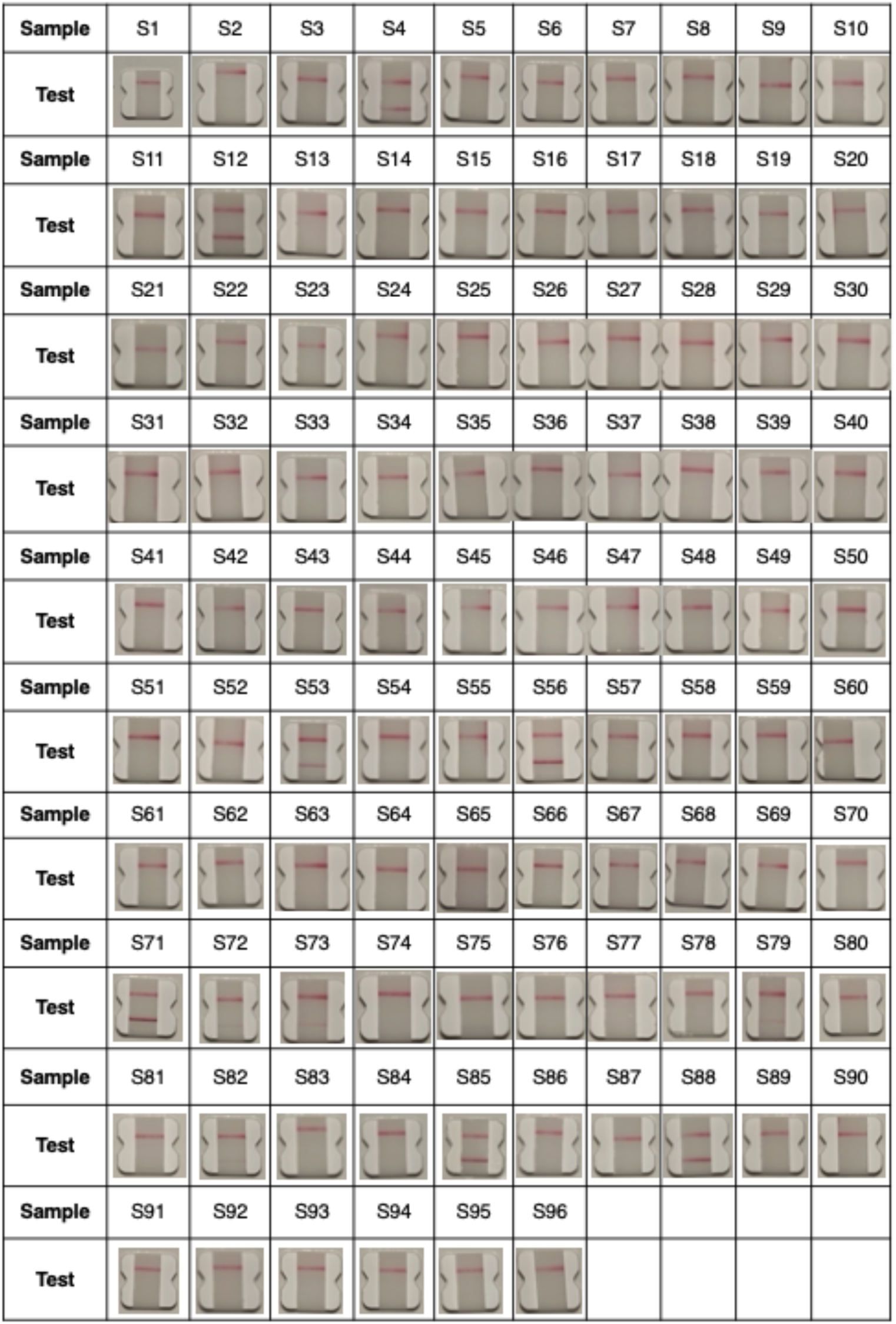
BinaxNow results on clinical samples. Images of positive and negative test results with BinaxNow on unextracted NP swab samples.

**Supplementary Fig. 10.**
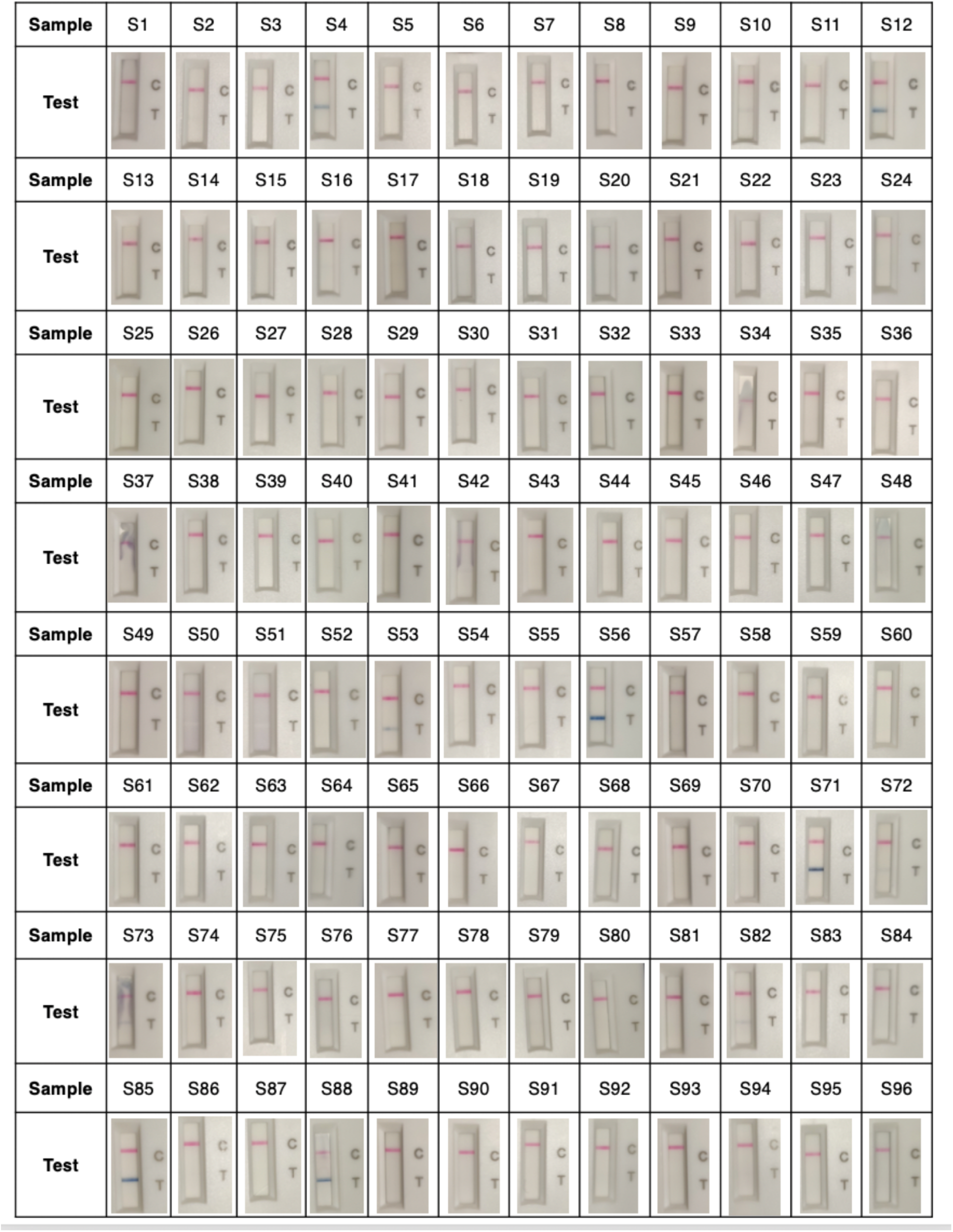
CareStart results on clinical samples. Images of positive and negative test results with CareStart on unextracted nasopharyngeal NP swab samples.

**Supplementary Fig. 11.**
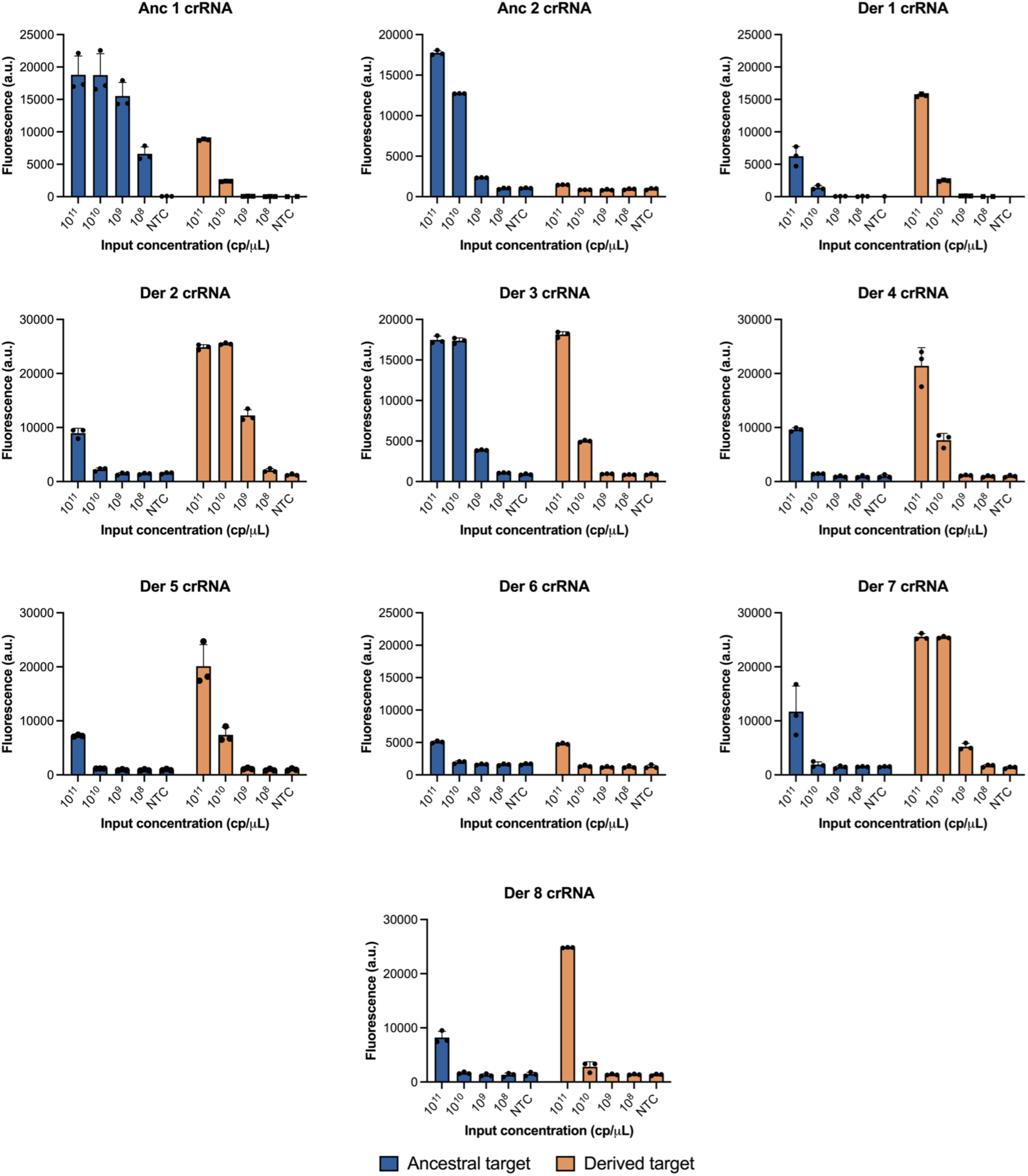
Selection of ancestral (anc) and derived (der) crRNAs for the S gene 69/70 deletion assay. Cas13a detection of ancestral and derived synthetic RNA targets with different anc and der crRNAs. NTC, no target control. Centre = mean and error bar = standard deviation (s.d.) for 3 technical replicates.

**Supplementary Fig. 12.**
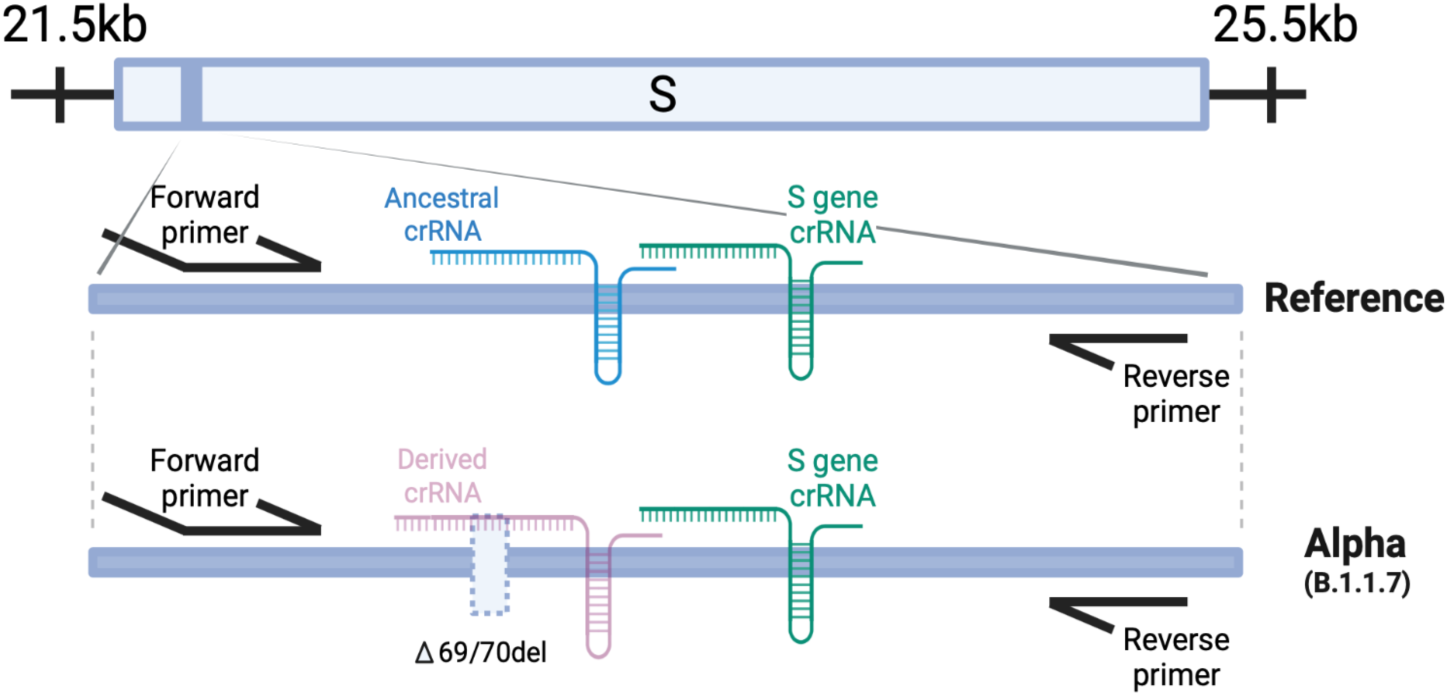
Location of the SHINEv2 assay for the 69/70 deletion detection. Schematic of location of the S gene and 69/70 deletion SHINEv2 assays within the S gene of SARS-CoV-2 for the reference strain and Alpha VOC. Dashed rectangle, deletion at amino acid positions 69 and 70 (Δ69/70del).

**Supplementary Fig. 13.**
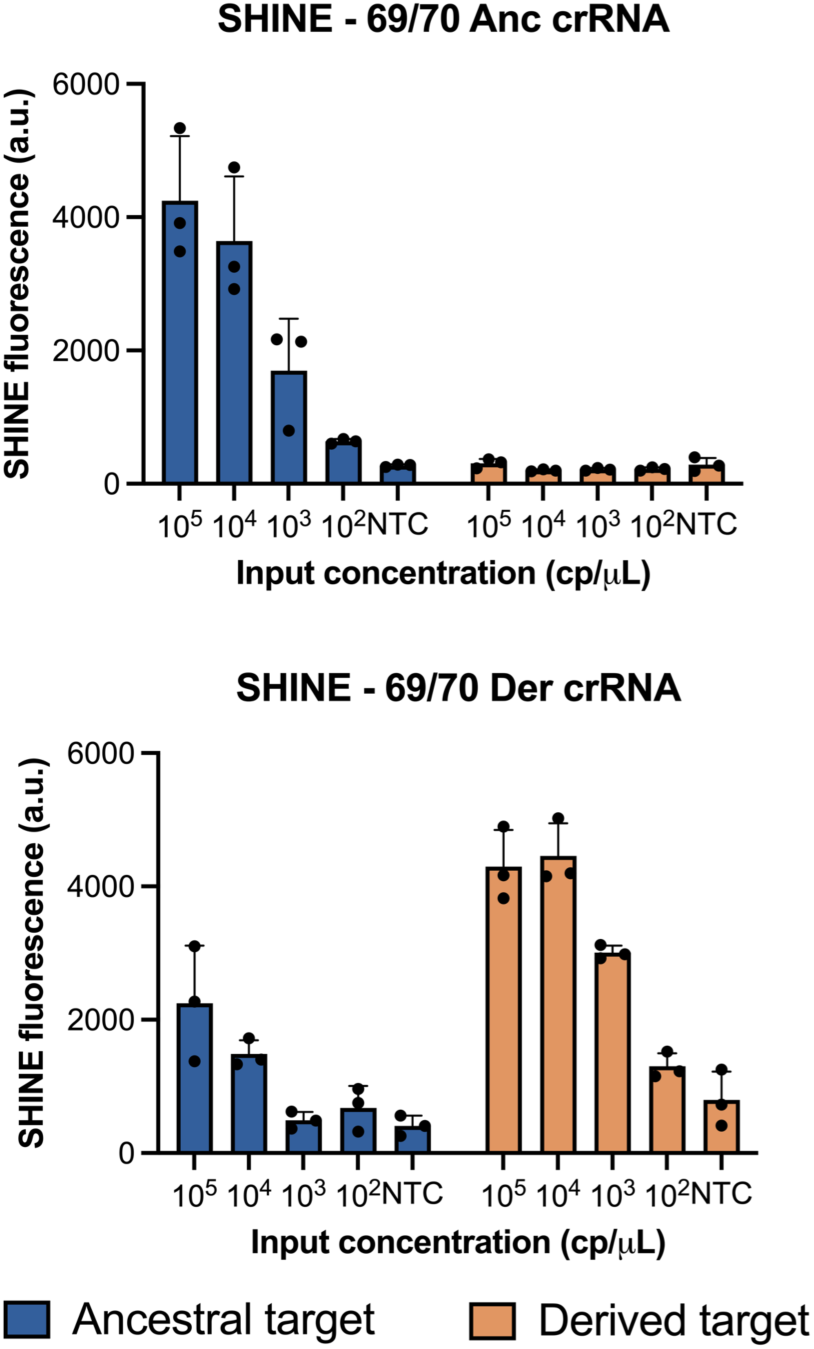
Performance of 69/70 deletion assay on synthetic RNA targets. SHINE fluorescence of the 69/70 ancestral (anc) and derived (der) assays on synthetic SARS-CoV-2 RNA targets after 90 minutes. NTC, no target control. Center = mean and error bars = s.d. for 3 technical replicates.

**Supplementary Fig. 14.**
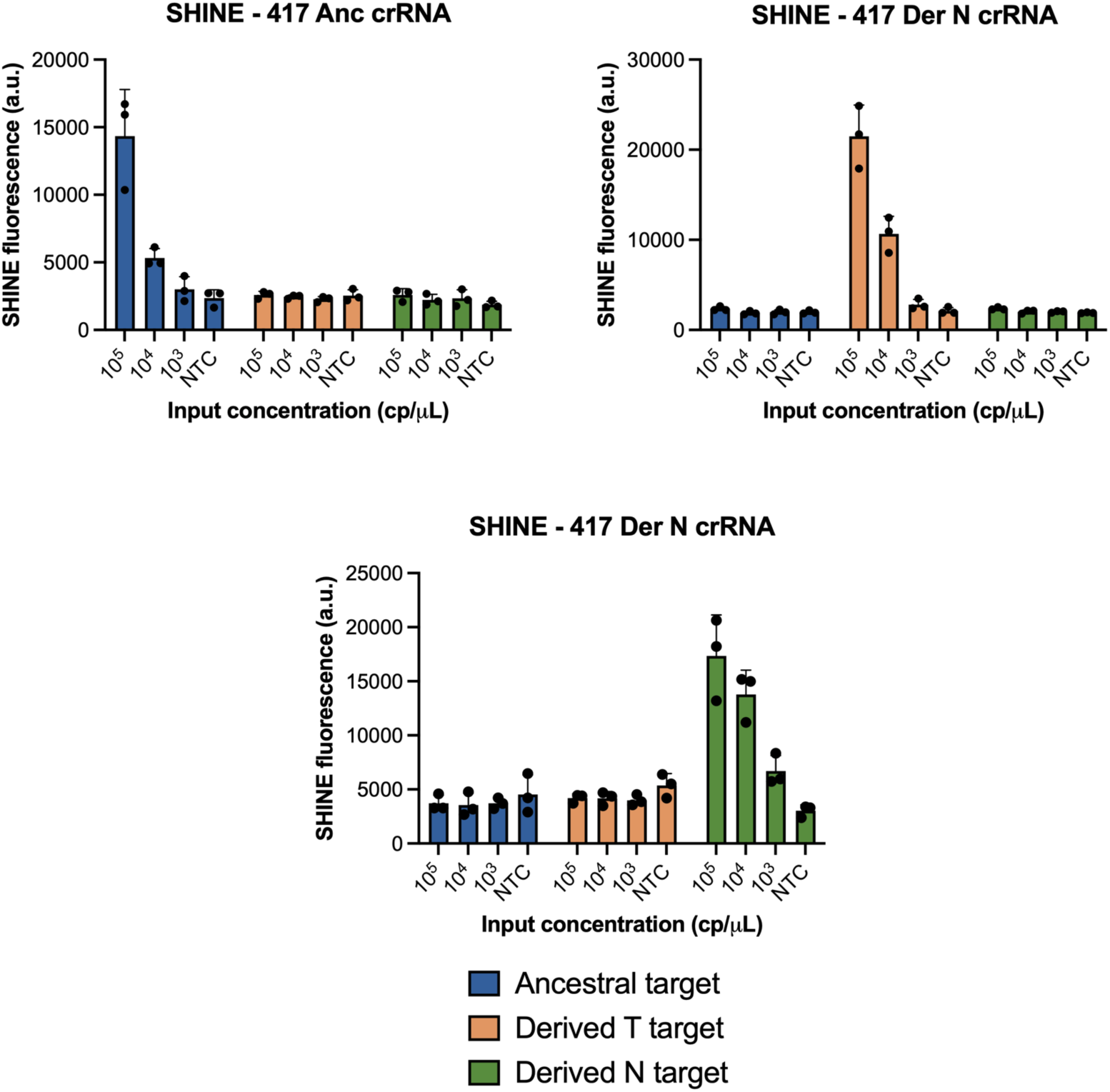
Performance of 417 SNP detection assay on synthetic RNA targets. SHINE fluorescence of the 417 ancestral (anc), derived T (der T) and derived N (der N) assays on synthetic SARS-CoV-2 RNA targets after 90 minutes. NTC, no target control. Center = mean and error bars = s.d. for 3 technical replicates.

**Supplementary Fig. 15.**
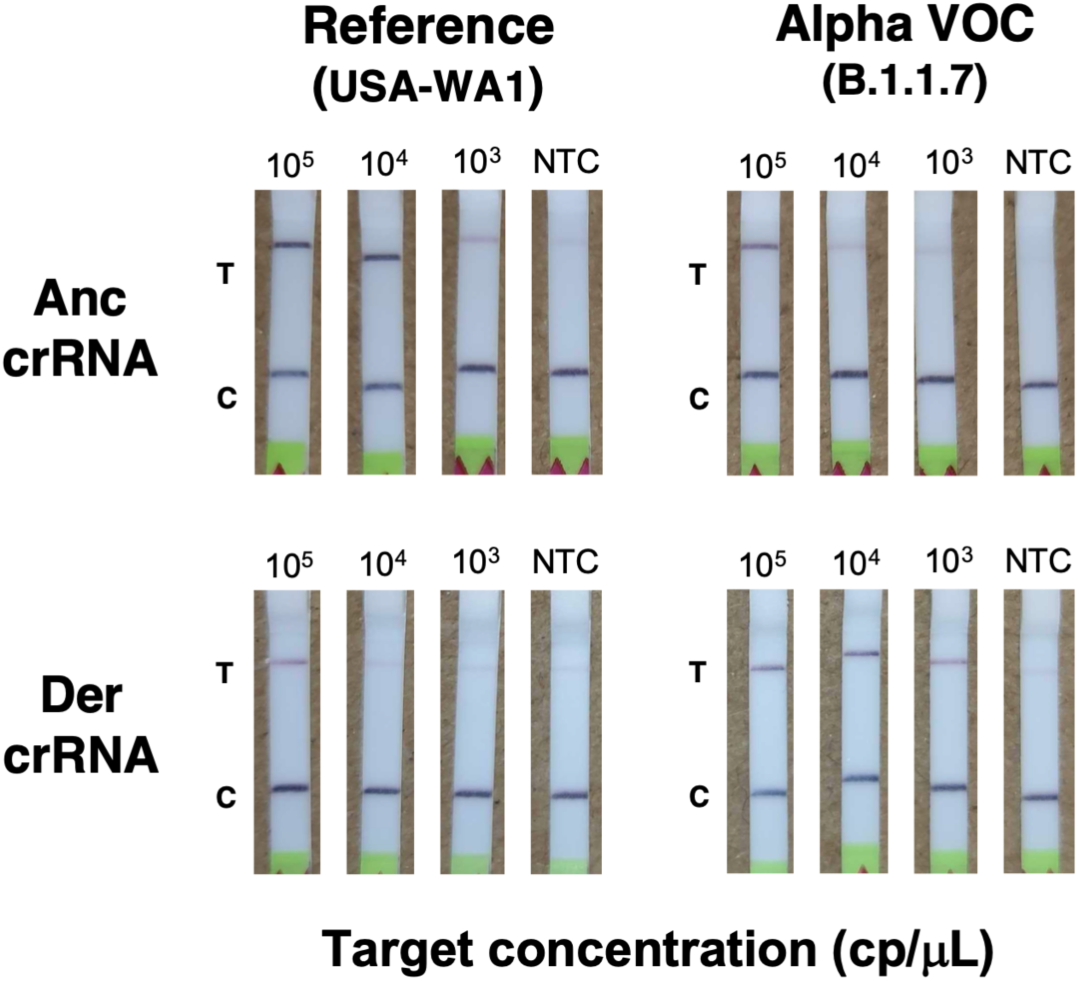
Lateral-flow detection of Alpha VOC in contrived clinical samples. Colorimetric lateral flow based detection of SARS-CoV-2 RNA in contrived clinical samples using the 69/70 SHINEv2 assay. SHINEv2 incubation time: 90 minutes. NTC, no-target control. T, test line; C, control line.

**Supplementary Fig. 16.**
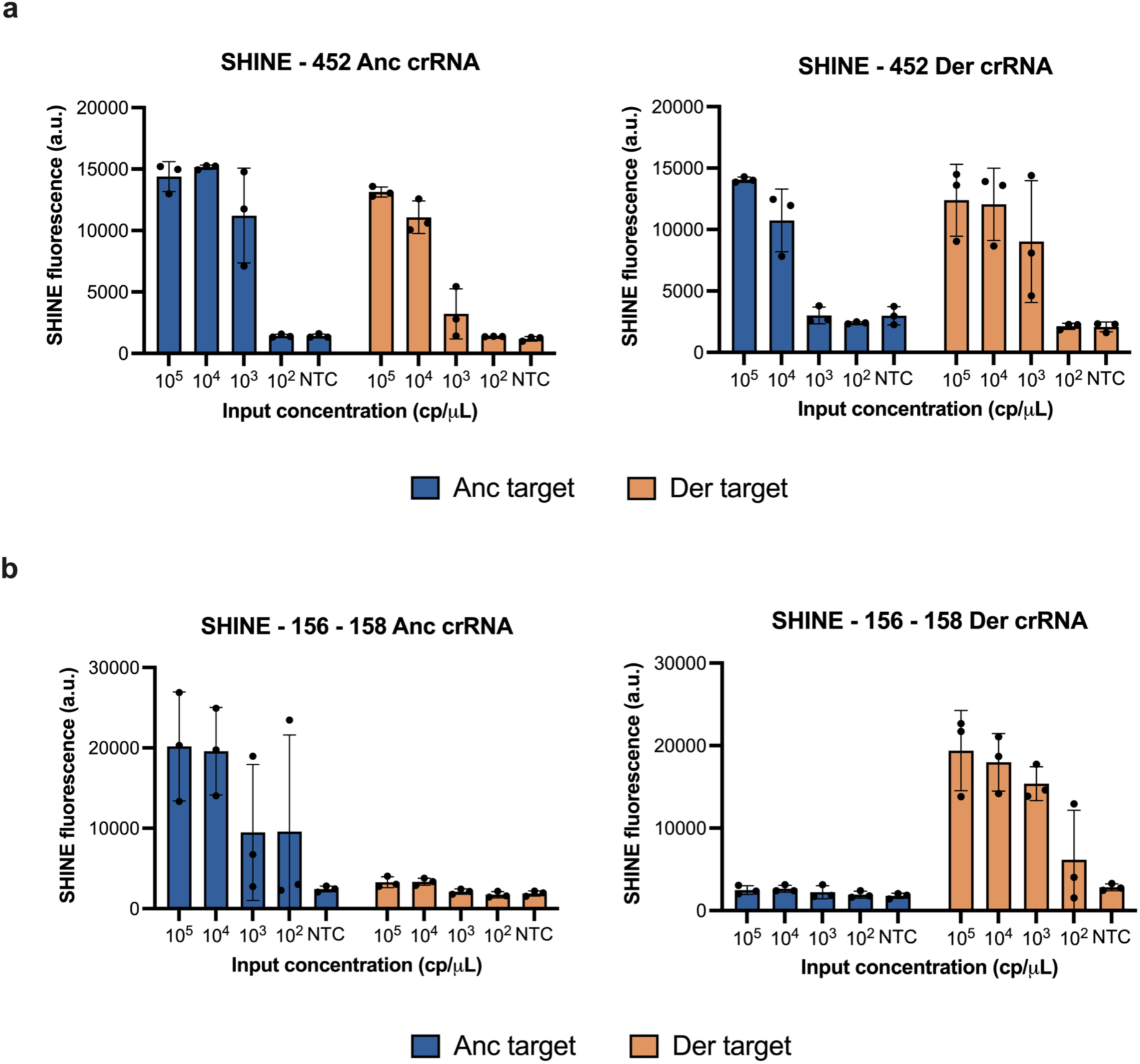
Performance of SARS-CoV-2 Delta assays on full genome synthetic RNA standards. **a,b.** SHINE fluorescence of the **(a)** 452 and **(b)** 156 - 158 ancestral (anc) and derived (der) assays on full genome synthetic RNA standards after 90 minutes. NTC, no target control. Center = mean and error bars = s.d. for 3 technical replicates.

**Supplementary Fig. 17.**
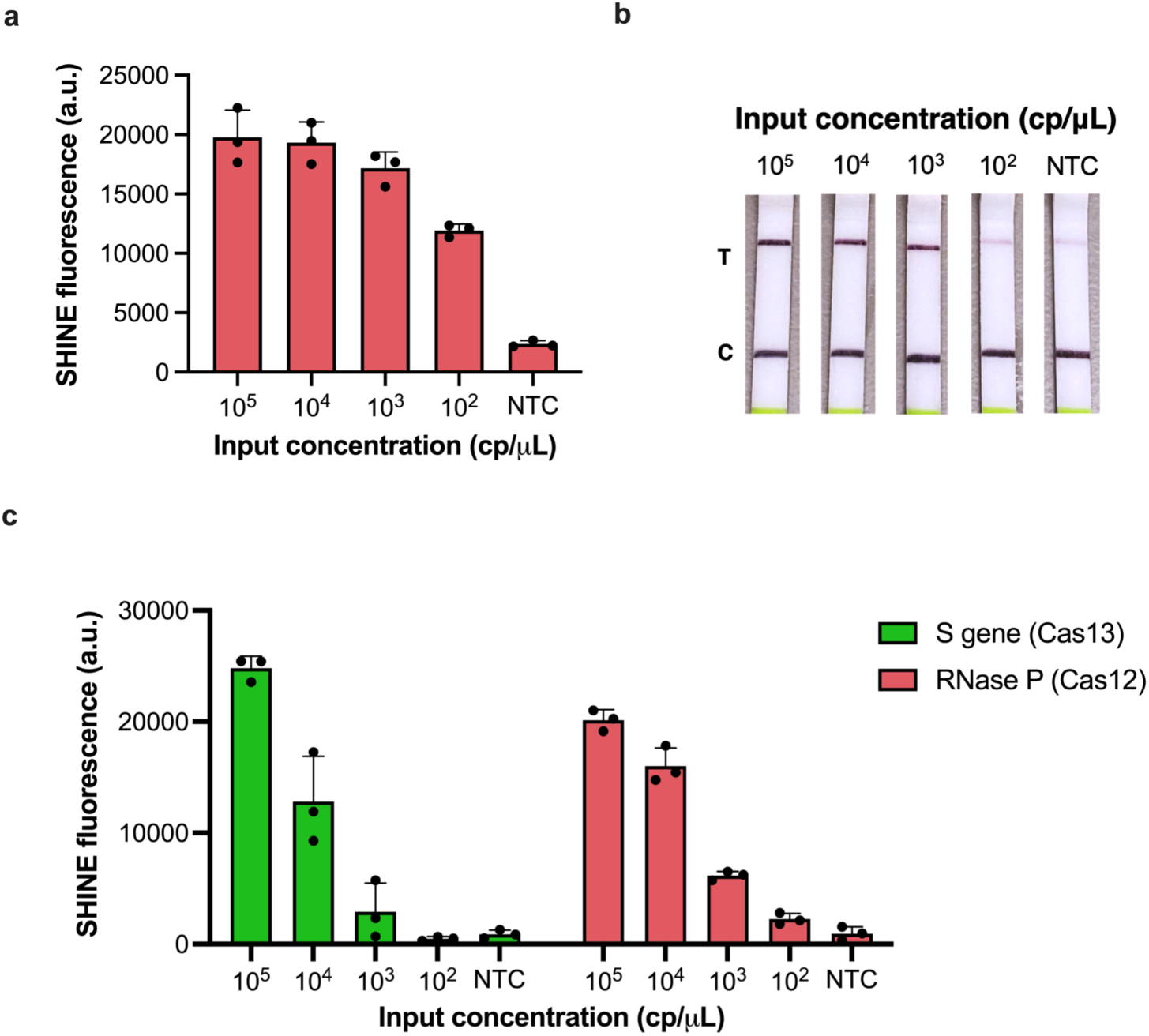
Development of Cas12-based SHINE assay for RNase P detection. **a.** SHINE fluorescence of the RNase P assay on synthetic DNA target after 90 minutes; NTC = no target control **b.** Lateral-flow detection of synthetic DNA target using Cas12-based SHINE assay, after 90 minute incubation; NTC = no target control C = control band; T = test band **c.** SHINE fluorescence of the duplex SARS-CoV-2 S gene and RNase P assays on synthetic nucleic acid targets after 90 minutes. NTC = no target control. For **a** and **c**, center = mean and error bars = s.d. for 3 technical replicates.

**Supplementary Fig. 18.**
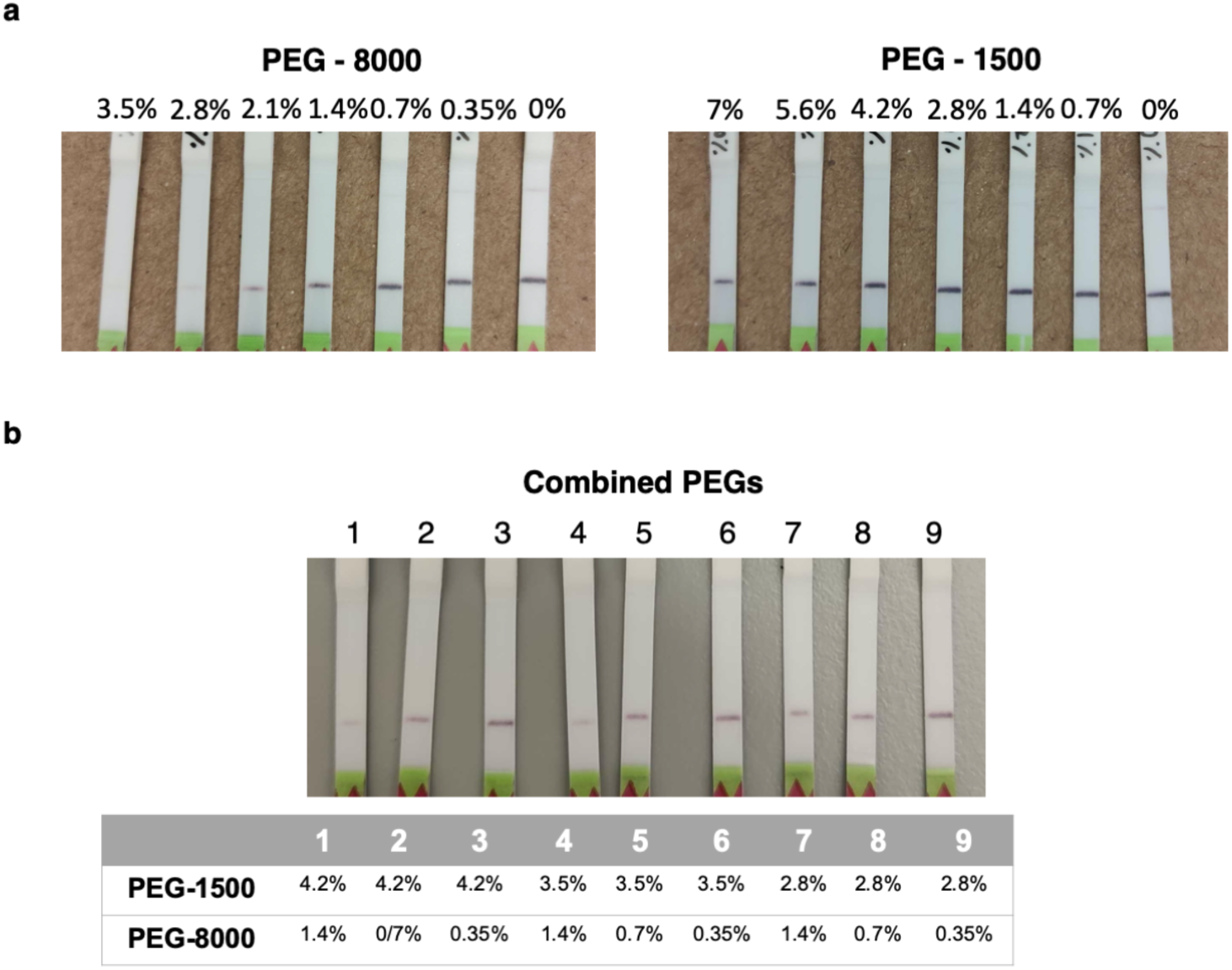
Effect of PEG on flow through the strip. **a** and **b,** images of lateral flow strips from SHINEv2 reactions with PEGs of different molecular weights and concentrations **(a)** separately and **(b)** in combination, in the absence of an RNA target.

**Supplementary Fig. 19.**
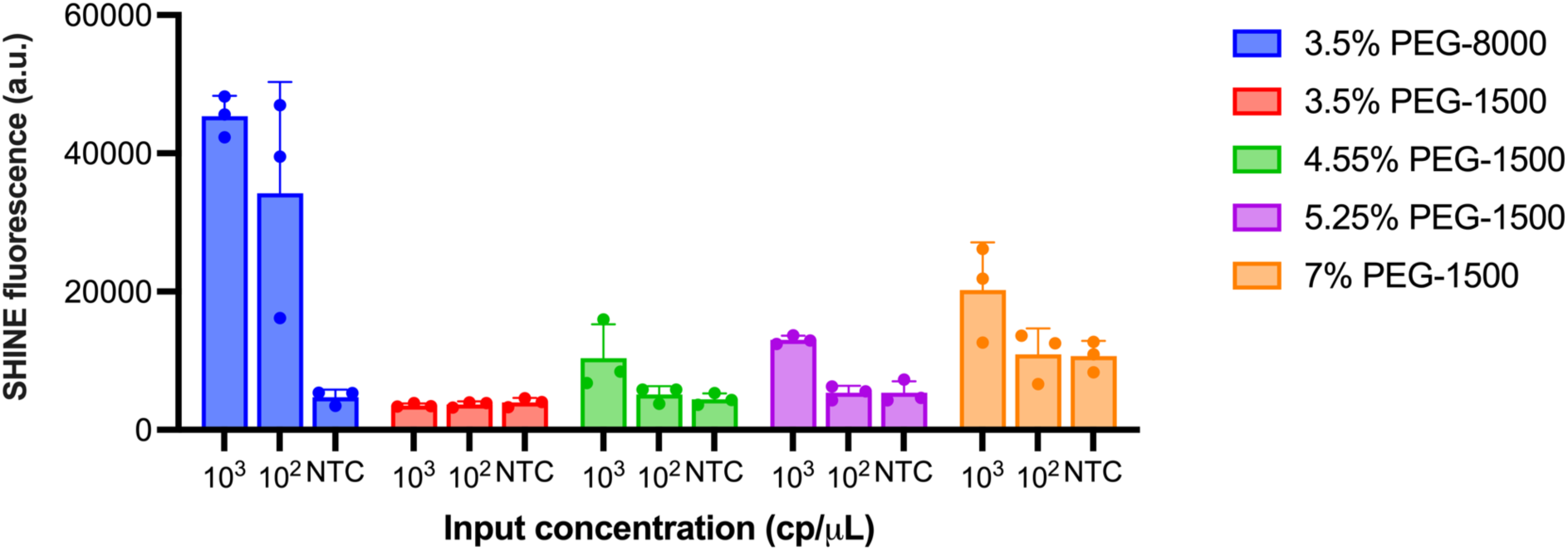
Effect of PEG concentration on SHINE performance. SHINE fluorescence on synthetic SARS-CoV-2 RNA targets (S gene) relative to PEG molecular weight and concentration. Reactions incubated for 90 minutes. NTC, no target control. Center = mean and error bars = s.d. for 3 technical replicates.

**Supplementary Fig. 20.**
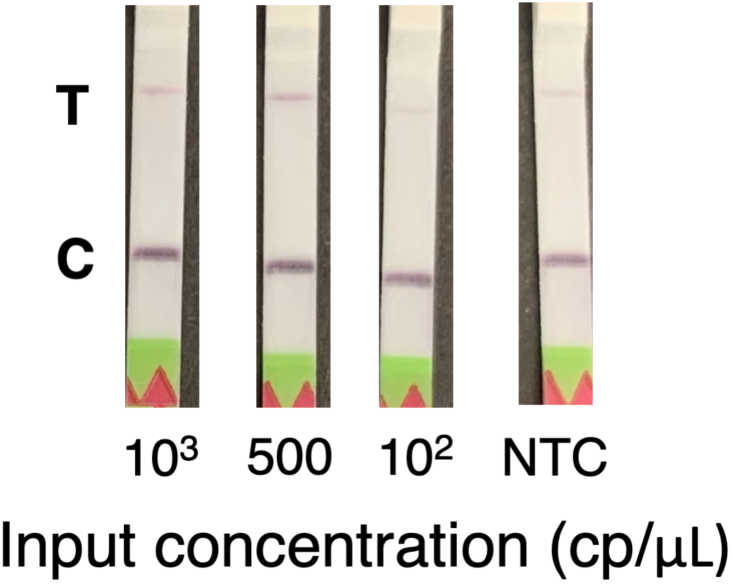
Performance of equipment-free SHINEv2 at room temperature. Lateral flow detection of full genome synthetic RNA standards in lysis solution-treated UTM using SHINEv2, after a 90 minute incubation at 25°C. C = control band; T = test band; NTC = no target control.

**Supplementary Fig. 21.**
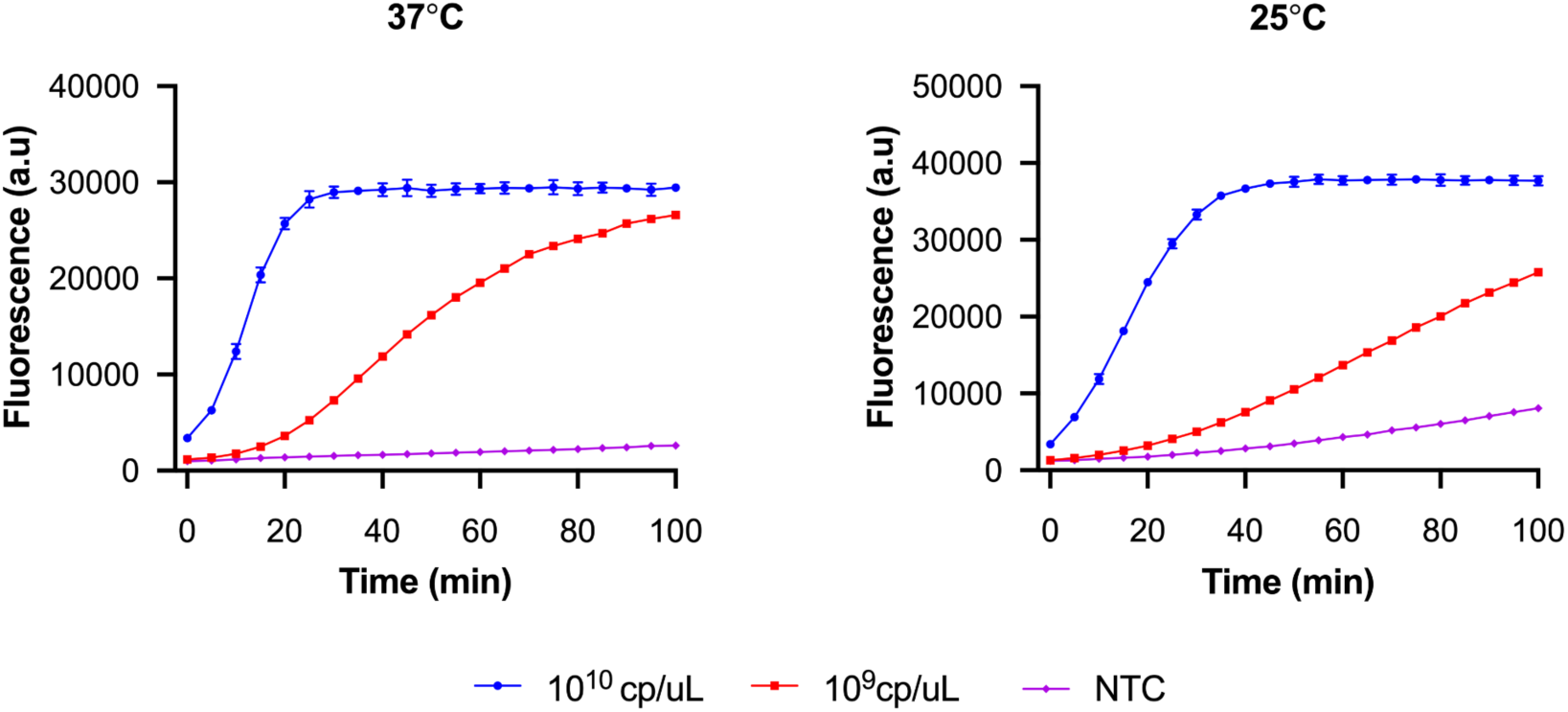
Temperature has a limited effect on Cas13-based detection. Cas13-detection of synthetic RNA target (S gene) at 37°C and 25°C. NTC, non target control. Centre = mean and error bar = standard deviation (s.d.) for 3 technical replicates.

**Supplementary Table 1.**
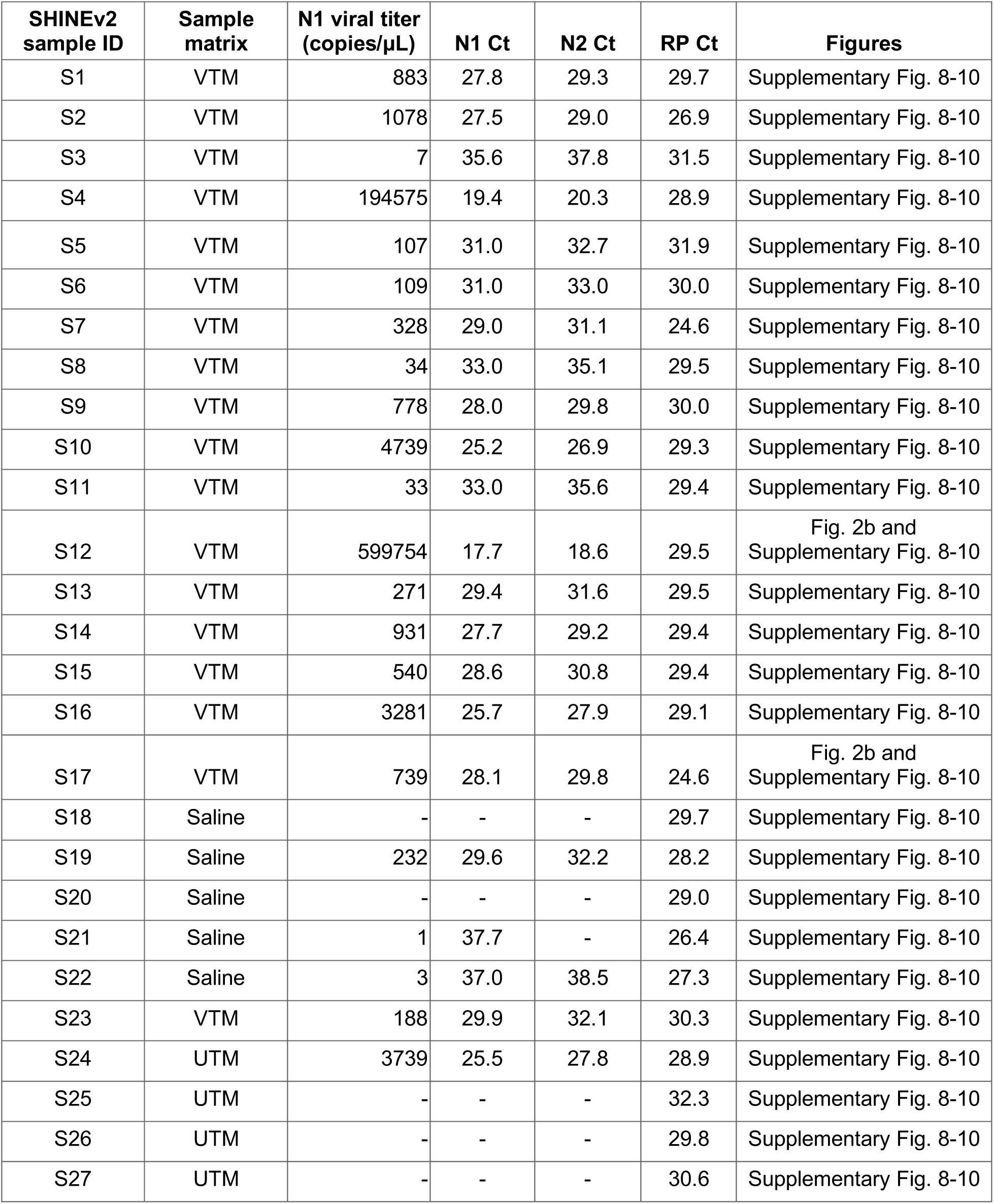
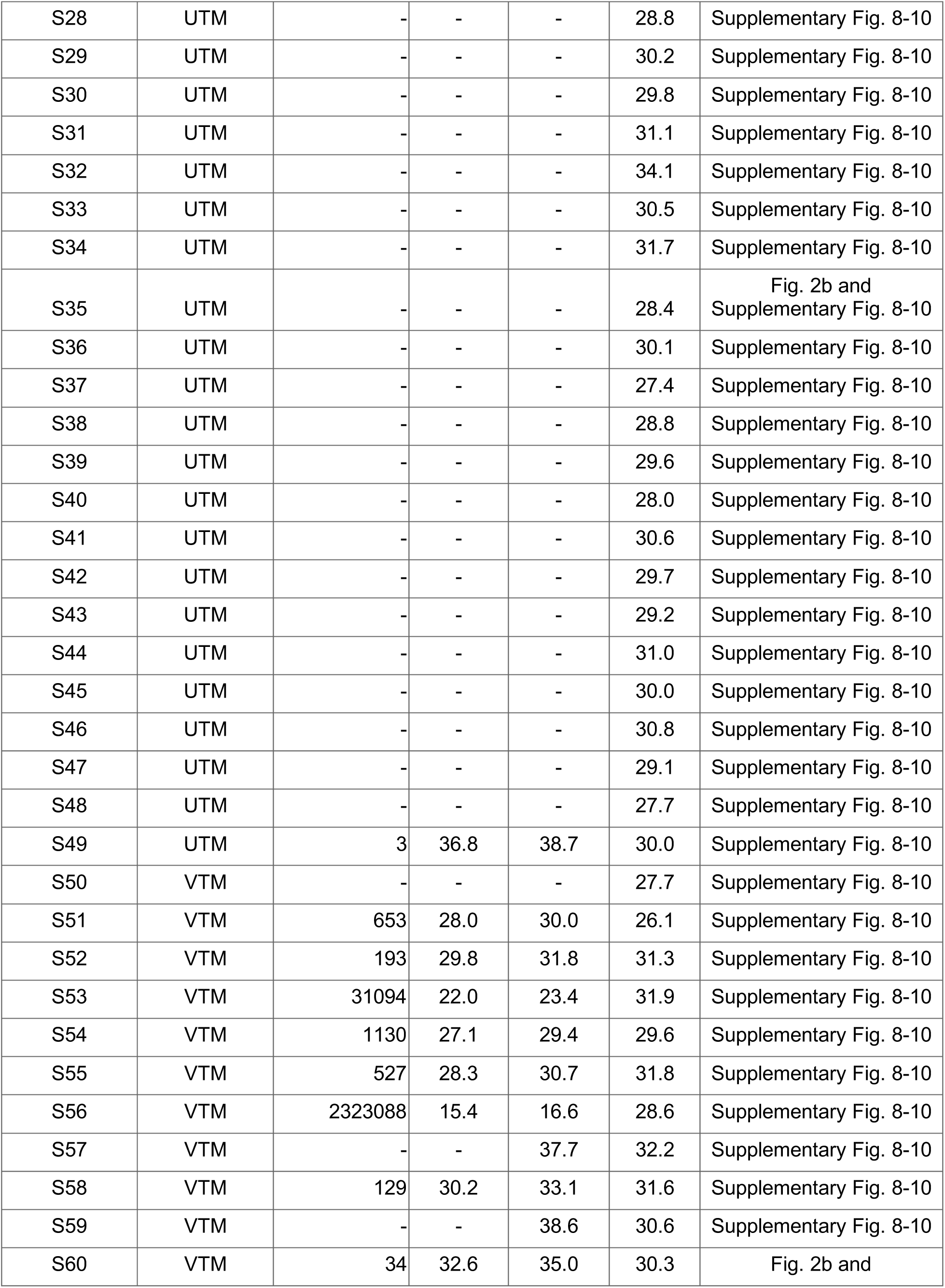
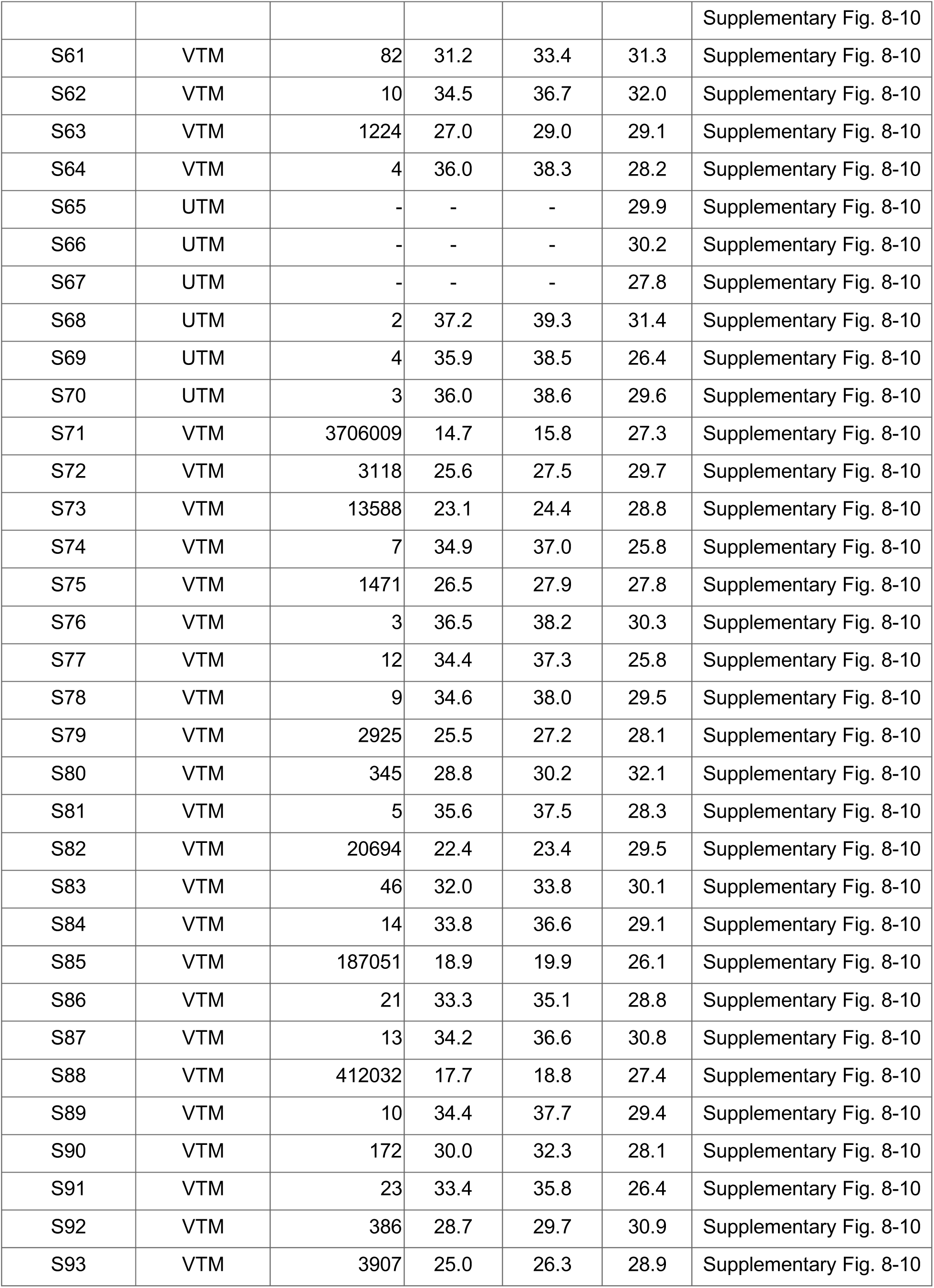
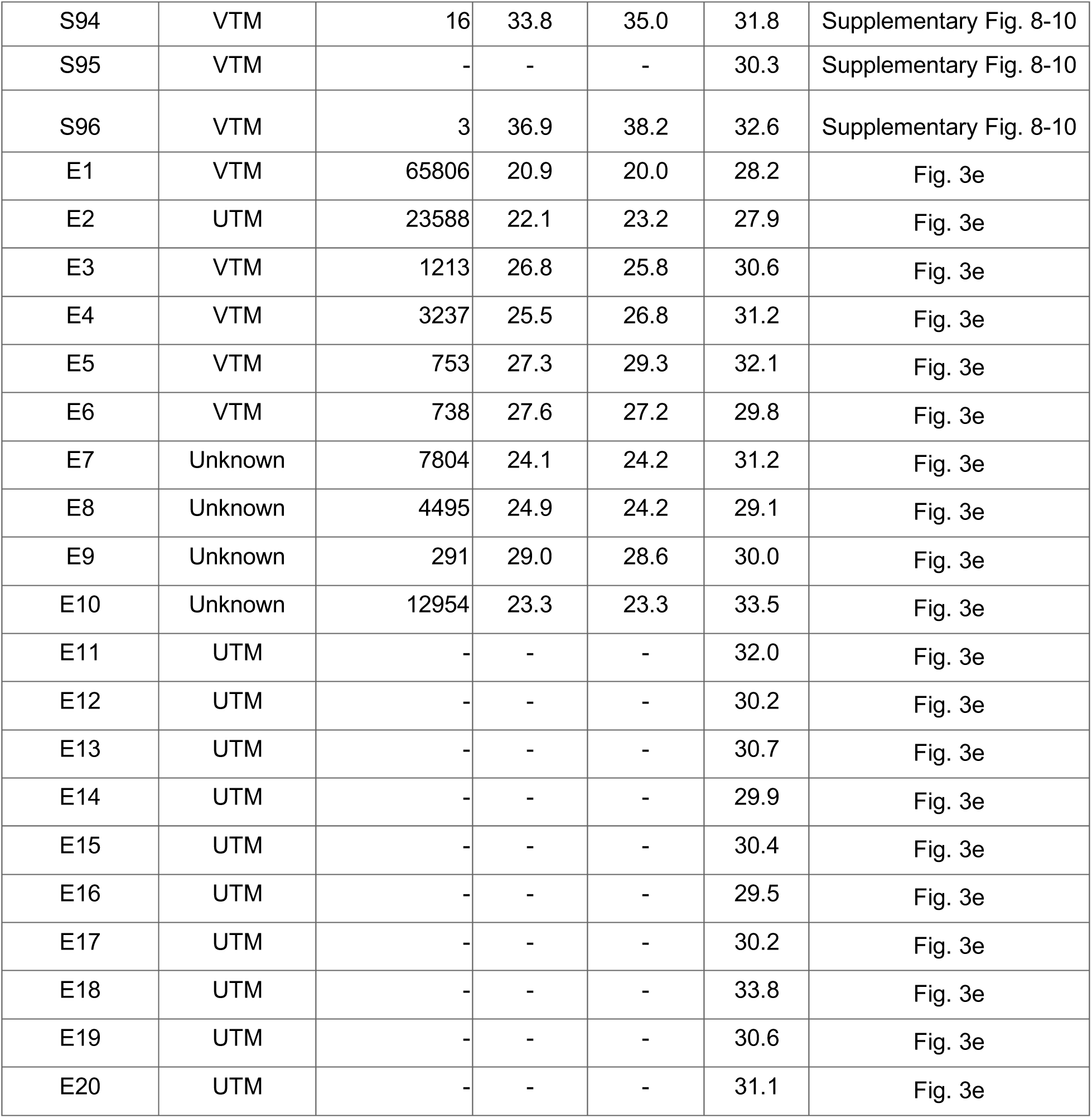
Patient sample information. Samples S1 - S96 were used without extraction, whereas RNA was extracted from samples E1 - E20 before use. See *methods* for details. UTM, universal viral transport medium; VTM, viral transport medium; RP, RNase P.

**Supplementary Table 2.**
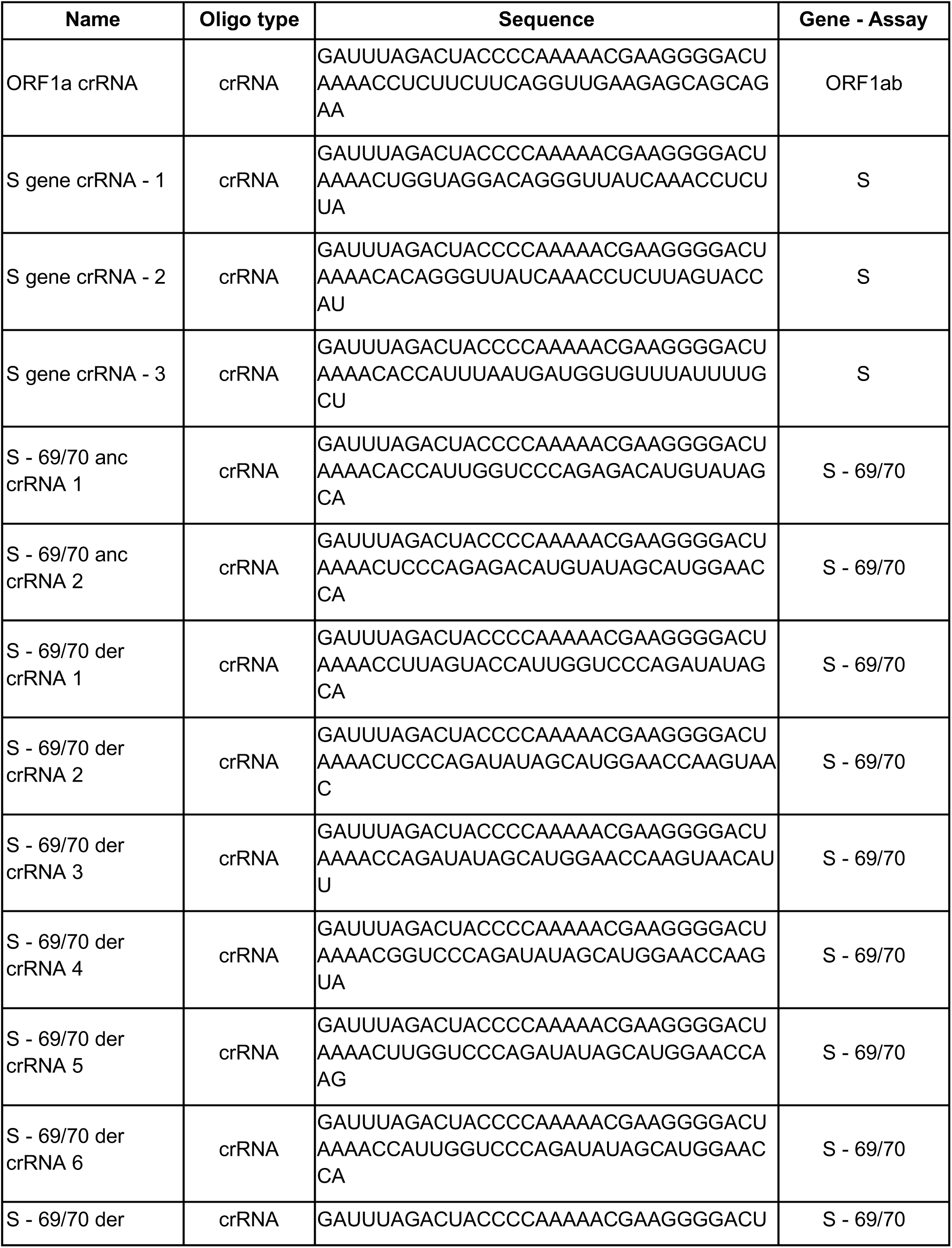
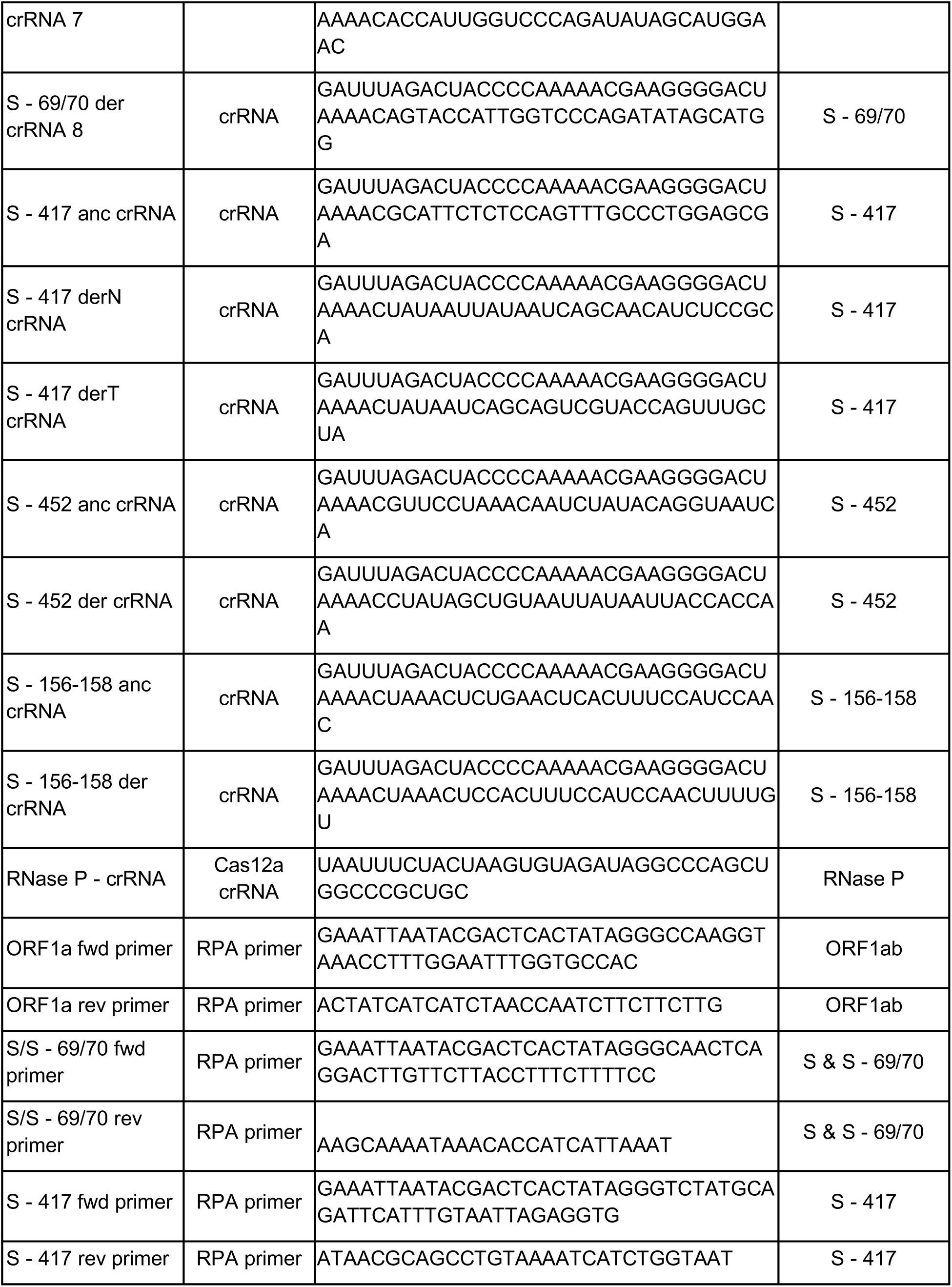
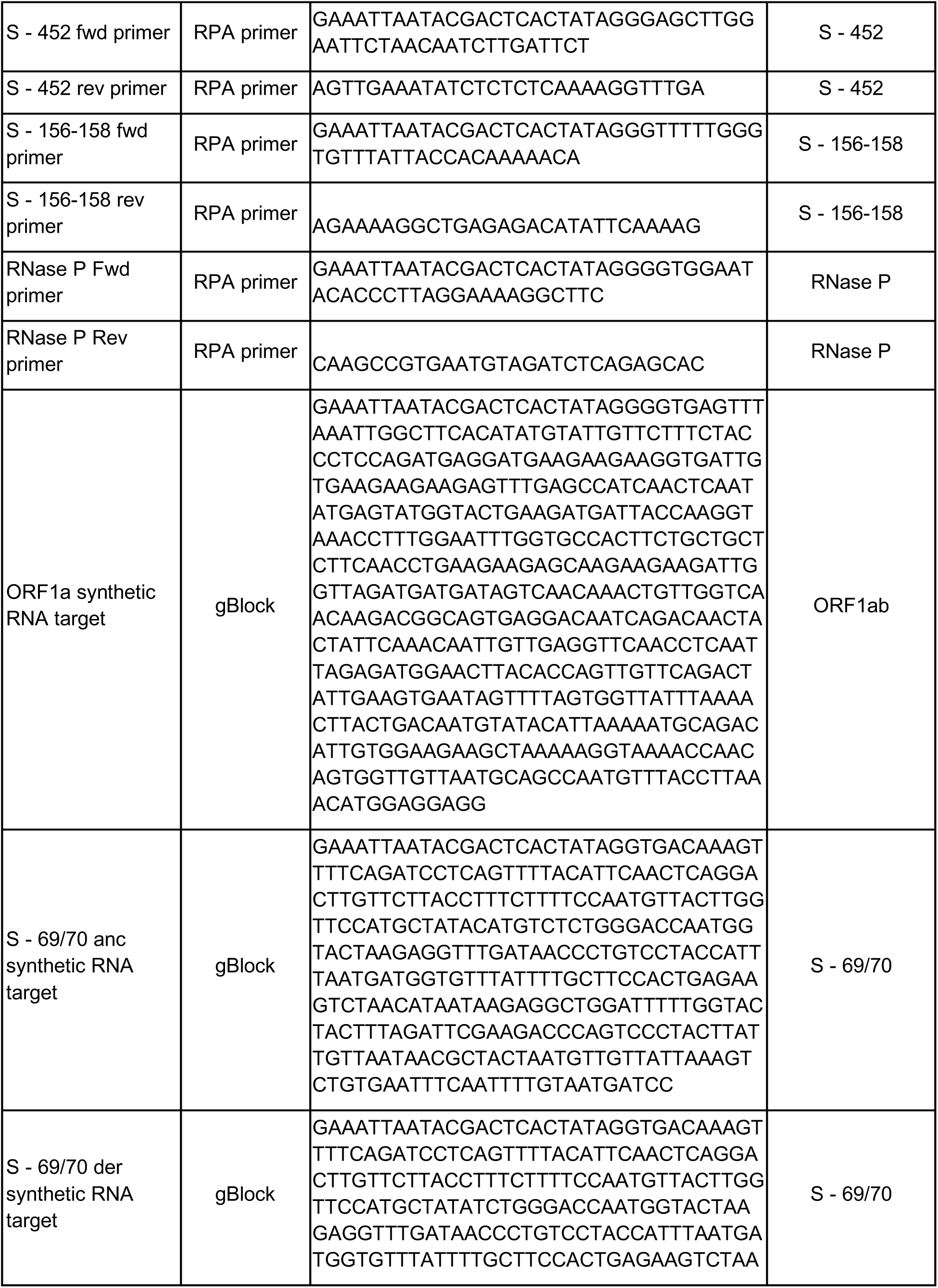
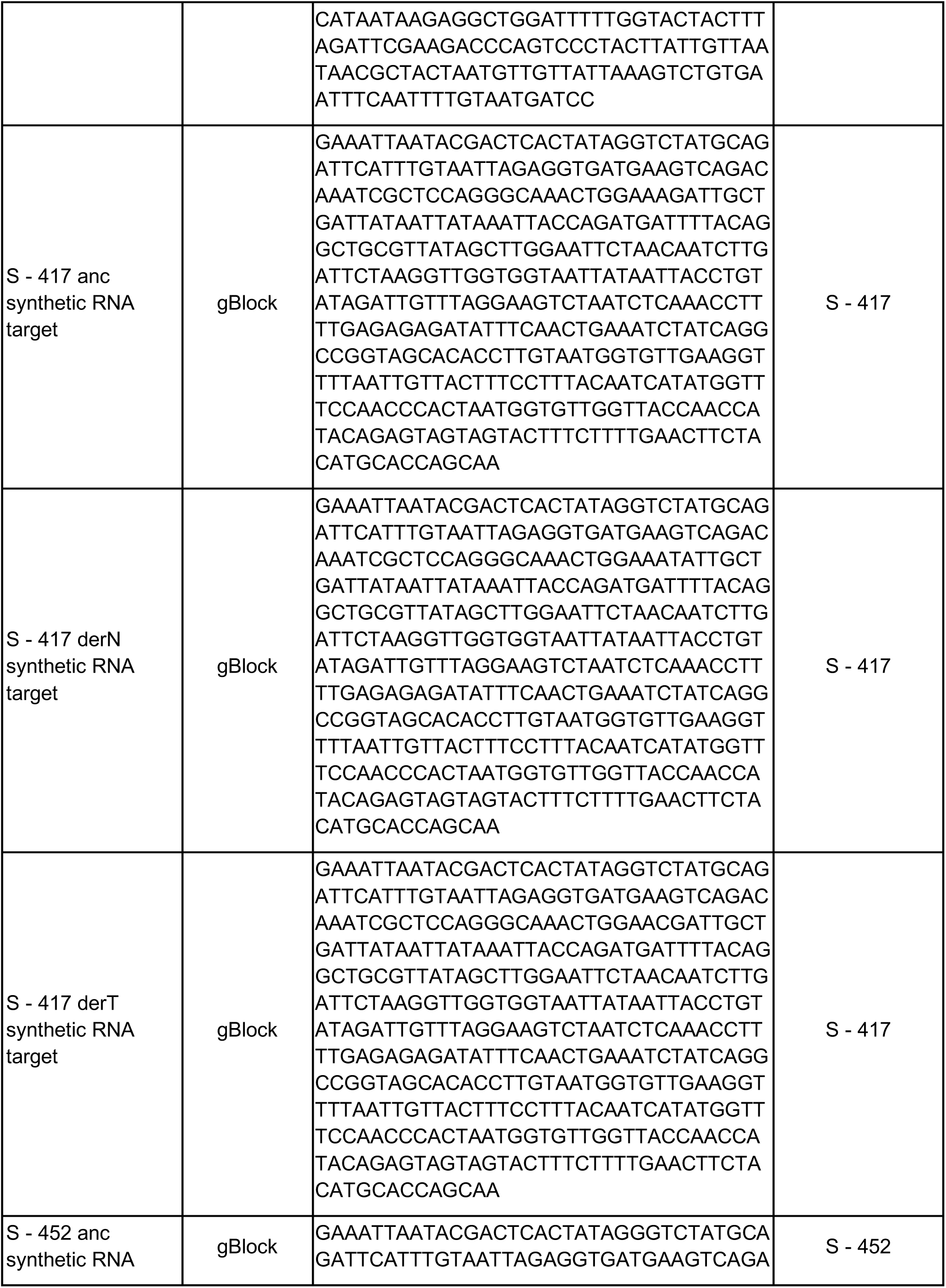
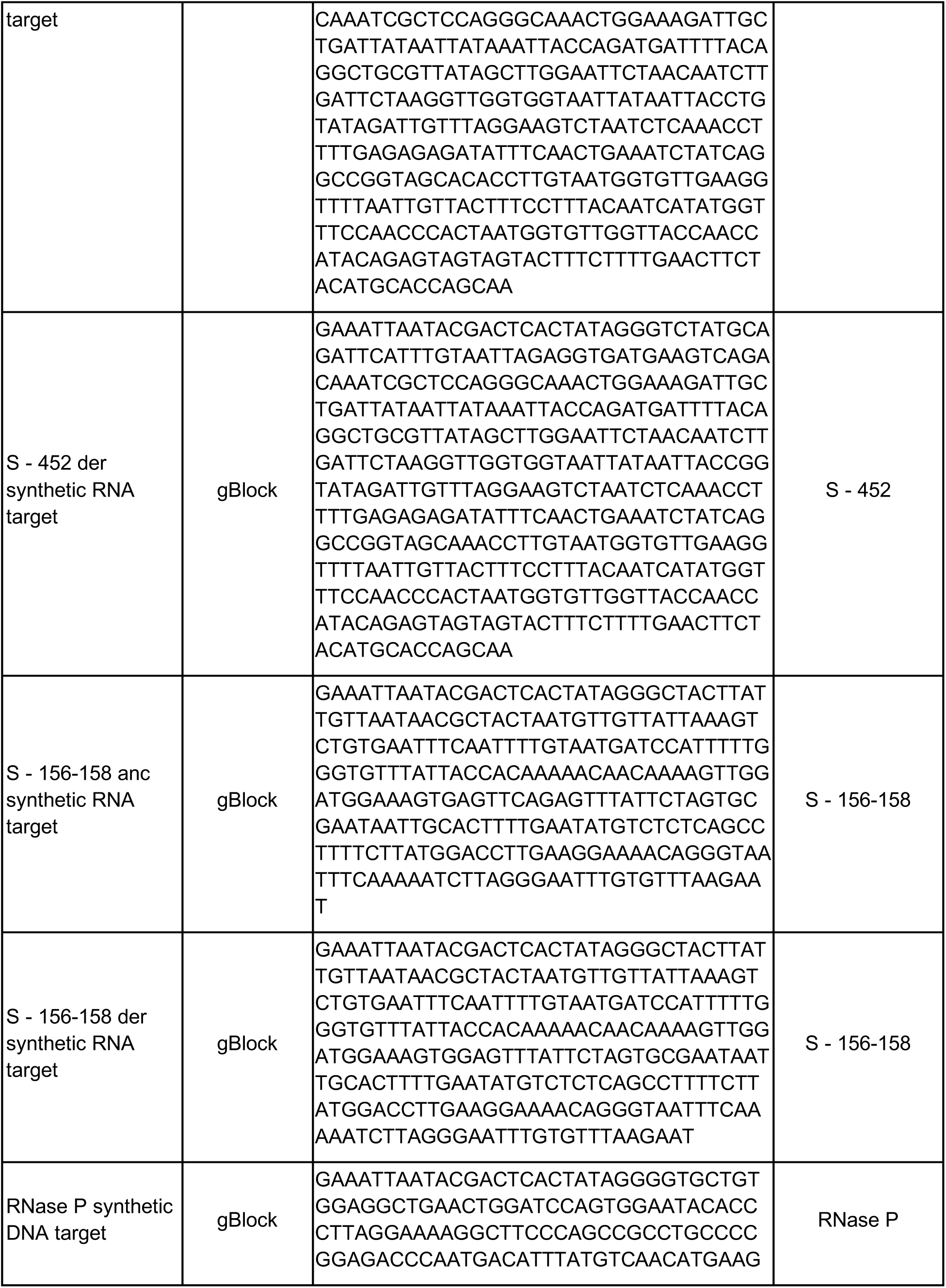
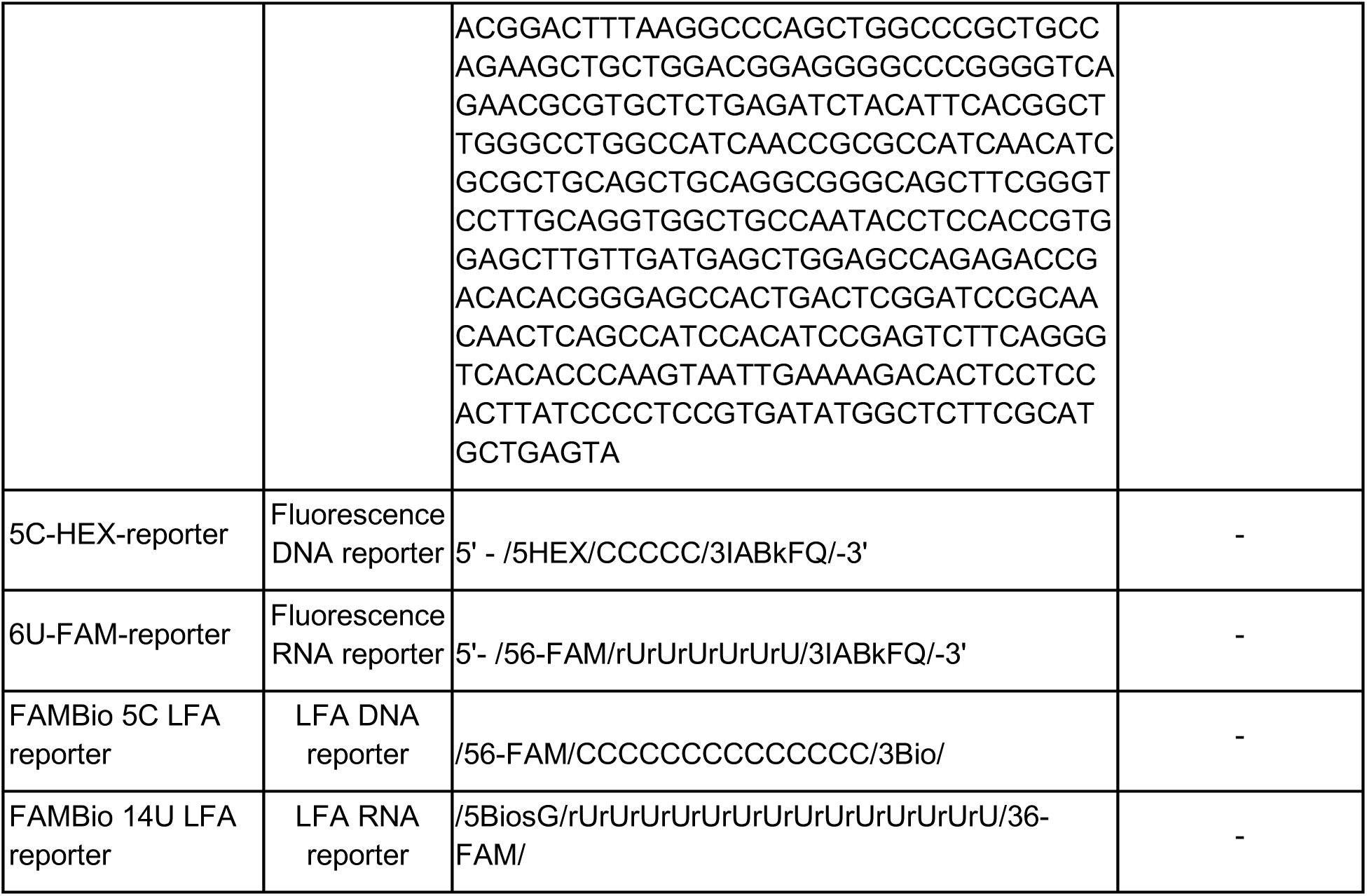
Oligonucleotides used in this study.

## References

1. Emerging COVID-19 success story: South Korea learned the lessons of MERS. https://ourworldindata.org/covid-exemplar-south-korea.

2. Summers, J. et al. Potential lessons from the Taiwan and New Zealand health responses to the COVID-19 pandemic. Lancet Reg Health West Pac 4, 100044 (2020).

3. Pavelka, M. et al. The impact of population-wide rapid antigen testing on SARS-CoV-2 prevalence in Slovakia. Science 372, 635–641 (2021).

4. Walensky, R. P. & del Rio, C. From Mitigation to Containment of the COVID-19 Pandemic. JAMA vol. 323 1889 (2020).

5. Mina, M. J. & Andersen, K. G. COVID-19 testing: One size does not fit all. Science 371, 126–127 (2021).

6. Mögling, R. et al. Delayed laboratory response to COVID-19 caused by molecular diagnostic contamination. Emerg. Infect. Dis. 26, 1944–1946 (2020).

7. McKay, S. L. et al. Performance Evaluation of Serial SARS-CoV-2 Rapid Antigen Testing During a Nursing Home Outbreak. Ann. Intern. Med. (2021) doi:10.7326/M21-0422.

8. Ferguson, J. et al. Validation testing to determine the sensitivity of lateral flow testing for asymptomatic SARS-CoV-2 detection in low prevalence settings: Testing frequency and public health messaging is key. PLoS Biol. 19, e3001216 (2021).

9. van Kampen, J. J. A. et al. Duration and key determinants of infectious virus shedding in hospitalized patients with coronavirus disease-2019 (COVID-19). Nat. Commun. 12, 267 (2021).

10. Pray, I. W., et al. Performance of an Antigen-Based Test for Asymptomatic and Symptomatic SARS-CoV-2 Testing at Two University Campuses - Wisconsin, September-October 2020. MMWR Morb. Mortal. Wkly. Rep. 69, 1642–1647 (2021).

11. Rabe, B. A. & Cepko, C. SARS-CoV-2 detection using isothermal amplification and a rapid, inexpensive protocol for sample inactivation and purification. Proc. Natl. Acad. Sci. U. S. A. 117, 24450–24458 (2020).

12. Thi, V. L. D. et al. A colorimetric RT-LAMP assay and LAMP-sequencing for detecting SARS-CoV-2 RNA in clinical samples. Sci. Transl. Med. 12, (2020).

13. Lucira™ COVID-19 Test Kit Instructions for Use. https://www.fda.gov/media/147494/download.

14. Land, K. J., Boeras, D. I., Chen, X.-S., Ramsay, A. R. & Peeling, R. W. REASSURED diagnostics to inform disease control strategies, strengthen health systems and improve patient outcomes. Nat Microbiol 4, 46–54 (2019).

15. Gootenberg, J. S. et al. Nucleic acid detection with CRISPR-Cas13a/C2c2. Science 356, 438–442 (2017).

16. Chen, J. S. et al. CRISPR-Cas12a target binding unleashes indiscriminate single-stranded DNase activity. Science 360, 436–439 (2018).

17. Li, S.-Y. et al. CRISPR-Cas12a-assisted nucleic acid detection. Cell Discov 4, 20 (2018).

18. Gootenberg, J. S. et al. Multiplexed and portable nucleic acid detection platform with Cas13, Cas12a, and Csm6. Science 360, 439–444 (2018).

19. Myhrvold, C. et al. Field-deployable viral diagnostics using CRISPR-Cas13. Science 360, 444–448 (2018).

20. Fozouni, P. et al. Amplification-free detection of SARS-CoV-2 with CRISPR-Cas13a and mobile phone microscopy. Cell 184, 323–333.e9 (2021).

21. Liu, T. Y. et al. Accelerated RNA detection using tandem CRISPR nucleases. medRxiv (2021) doi:10.1101/2021.03.19.21253328.

22. Joung, J. et al. Detection of SARS-CoV-2 with SHERLOCK One-Pot Testing. N. Engl. J. Med. 383, 1492–1494 (2020).

23. Arizti-Sanz, J., et al. Streamlined inactivation, amplification, and Cas13-based detection of SARS-CoV-2. Nat. Commun. 11, 5921 (2020).

24. Harvey, W. T. et al. SARS-CoV-2 variants, spike mutations and immune escape. Nat. Rev. Microbiol. 19, 409–424 (2021).

25. Konings, F. et al. SARS-CoV-2 Variants of Interest and Concern naming scheme conducive for global discourse. Nat Microbiol 6, 821–823 (2021).

26. Volz, E. et al. Assessing transmissibility of SARS-CoV-2 lineage B.1.1.7 in England. Nature 593, 266–269 (2021).

27. Lemieux, J. E. et al. Phylogenetic analysis of SARS-CoV-2 in Boston highlights the impact of superspreading events. Science 371, (2021).

28. Borges, V. et al. Tracking SARS-CoV-2 lineage B.1.1.7 dissemination: insights from nationwide spike gene target failure (SGTF) and spike gene late detection (SGTL) data, Portugal, week 49 2020 to week 3 2021. Euro Surveill. 26, (2021).

29. Vogels, C. B. F. et al. Multiplex qPCR discriminates variants of concern to enhance global surveillance of SARS-CoV-2. PLoS Biol. 19, e3001236 (2021).

30. Brito, A. F. et al. Global disparities in SARS-CoV-2 genomic surveillance. (2021) doi:10.1101/2021.08.21.21262393.

31. Ackerman, C. M. et al. Massively multiplexed nucleic acid detection with Cas13. Nature vol. 582 277–282 (2020).

32. Casati, B. et al. ADESSO detects SARS-CoV-2 and its variants: extensive clinical validation of an optimised CRISPR-Cas13-based COVID-19 test. doi:10.1101/2021.06.17.21258371.

33. Jiao, C. et al. Noncanonical crRNAs derived from host transcripts enable multiplexable RNA detection by Cas9. Science 372, 941–948 (2021).

34. Chakraborty, D., Agrawal, A. & Maiti, S. Rapid identification and tracking of SARS-CoV-2 variants of concern. The Lancet vol. 397 1346–1347 (2021).

35. de Puig, H. et al. Minimally instrumented SHERLOCK (miSHERLOCK) for CRISPR-based point-of-care diagnosis of SARS-CoV-2 and emerging variants. Sci Adv 7, (2021).

36. Metsky, H. C. et al. Designing viral diagnostics with model-based optimization. bioRxiv 2020.11.28.401877 (2021) doi:10.1101/2020.11.28.401877.

37. Qian, J. et al. An enhanced isothermal amplification assay for viral detection. Nat. Commun. 11, 5920 (2020).

38. Policy for Coronavirus Disease-2019 Tests During the Public Health Emergency. https://www.fda.gov/media/135659/download.

39. Perchetti, G. A., Huang, M.-L., Mills, M. G., Jerome, K. R. & Greninger, A. L. Analytical Sensitivity of the Abbott BinaxNOW COVID-19 Ag Card. J. Clin. Microbiol. 59, (2021).

40. Xun, G., Lane, S. T., Petrov, V. A., Pepa, B. E. & Zhao, H. A rapid, accurate, scalable, and portable testing system for COVID-19 diagnosis. Nat. Commun. 12, 2905 (2021).

41. CDC 2019-Novel Coronavirus (2019-nCoV) Real-Time RT-PCR Diagnostic Panel. https://www.fda.gov/media/134922/download.

42. Pollock, N. R. et al. Performance and Implementation Evaluation of the Abbott BinaxNOW Rapid Antigen Test in a High-throughput Drive-through Community Testing Site in Massachusetts. doi:10.1101/2021.01.09.21249499.

43. Pollock, N. R. et al. Performance and Operational Evaluation of the Access Bio CareStart Rapid Antigen Test in a High-throughput Drive-through Community Testing Site in Massachusetts. doi:10.1101/2021.03.07.21253101.

44. Allan-Blitz, L.-T. & Klausner, J. D. A Real-World Comparison of SARS-CoV-2 Rapid Antigen vs. Polymerase Chain Reaction Testing in Florida. J. Clin. Microbiol. JCM0110721 (2021) doi:10.1128/JCM.01107-21.

45. Okoye, N. C. et al. Performance Characteristics of BinaxNOW COVID-19 Antigen Card for Screening Asymptomatic Individuals in a University Setting. J. Clin. Microbiol. 59, (2021).

46. Paltiel, A. D., Zheng, A. & Walensky, R. P. Assessment of SARS-CoV-2 Screening Strategies to Permit the Safe Reopening of College Campuses in the United States. JAMA Netw Open 3, e2016818 (2020).

47. Hodcroft, E. B. CoVariants: SARS-CoV-2 Mutations and Variants of Interest. https://covariants.org/ (2021).

48. Planas, D. et al. Reduced sensitivity of SARS-CoV-2 variant Delta to antibody neutralization. Nature 596, 276–280 (2021).

49. Crannell, Z. A., Rohrman, B. & Richards-Kortum, R. Equipment-free incubation of recombinase polymerase amplification reactions using body heat. PLoS One 9, e112146 (2014).

50. Evaluation of Isothermal Recombinase Polymerase Amplification Incubation Temperature Tolerance. Biotechniques 61, 328 (2016).

51. Yuan, C.-Q. et al. Universal and naked-eye gene detection platform based on CRISPR/Cas12a/13a system. doi:10.1101/615724.

52. Carter, J. G. et al. Ultrarapid detection of SARS-CoV-2 RNA using a reverse transcription–free exponential amplification reaction, RTF-EXPAR. Proc. Natl. Acad. Sci. U. S. A. 118, (2021).

53. Elbe, S. & Buckland-Merrett, G. Data, disease and diplomacy: GISAID’s innovative contribution to global health. Glob Chall 1, 33–46 (2017).

54. Shu, Y. & McCauley, J. GISAID: Global initiative on sharing all influenza data – from vision to reality. Euro Surveill. 22, (2017).

55. Mantena. In preparation.

56. BinaxNOW COVID-19 Antigen Self TEST. https://www.fda.gov/media/147254/download.

57. CareStart COVID-19 Antigen Rapid Diagnostic Test for the Detection of SARS-CoV-2 Antigen. https://www.fda.gov/media/142919/download.

